# Physiology-Informed Digital Twin-AI Framework Predicts Pacing Therapy Response in HFpEF

**DOI:** 10.64898/2026.03.06.26347199

**Authors:** Feng Gu, Margret Infeld, Noah A Schenk, Haolin Wan, Mothieshwar J Krishnan, Jamie A Cyr, Victoria E Sturgess, Emily Wittrup, Filip Ježek, Brian E Carlson, Tim van Loon, Xinwei Hua, Yi-Da Tang, Najarian Kayvan, Scott L Hummel, Joost Lumens, Markus Meyer, Daniel A Beard

## Abstract

**Background and Aims:** Heart failure with preserved ejection fraction (HFpEF) exhibits profound phenotypic heterogeneity, which likely contributes to variable therapeutic response. We developed a physiology-informed digital twin–AI framework to predict individual hemodynamic and myocardial energetic responses to accelerated atrial pacing and tested whether simulated physiologic response corresponds to responders in the myPACE randomized clinical trial.

**Methods:** Patient-specific digital twins were constructed for 146 HFpEF patients and used to train a variational autoencoder that generated a virtual HFpEF population (n = 25,000). The model simulated pacing-induced changes in left atrial pressure (LAP), systolic blood pressure (SBP), cardiac output (CO), and cardiac efficiency (CE; derived from myocardial oxygen-demand estimates). These simulations served as labels to train classifiers based on clinical variables available in myPACE, allowing us to examine associations with clinical end points and test a hypothesized relationship between CE and treatment response.

**Results:** Simulations revealed heterogeneous physiological responses, with 95.6% of virtual patients showing reduced LAP, 47.0% an SBP reduction greater than 8.5 mmHg, 93.8% increased CO, and 36.1% improved CE. Classifiers reproduced these patterns with high fidelity. In the myPACE trial, patients classified as having CE improvement or a larger SBP reduction experienced significantly greater 1-month improvements in quality-of-life scores and larger NT-proBNP reductions.

**Conclusions:** A physiology-informed digital twin–AI framework can predict hemodynamic and energetic responses corresponding to clinical benefit in HFpEF patients receiving accelerated atrial pacing. CE improvement functioned as a mechanistic indicator, while SBP reduction served as an accessible clinical correlate, offering mechanistically grounded guidance for patient-specific pacing and motivating prospective validation.

**Graphical Abstract:** 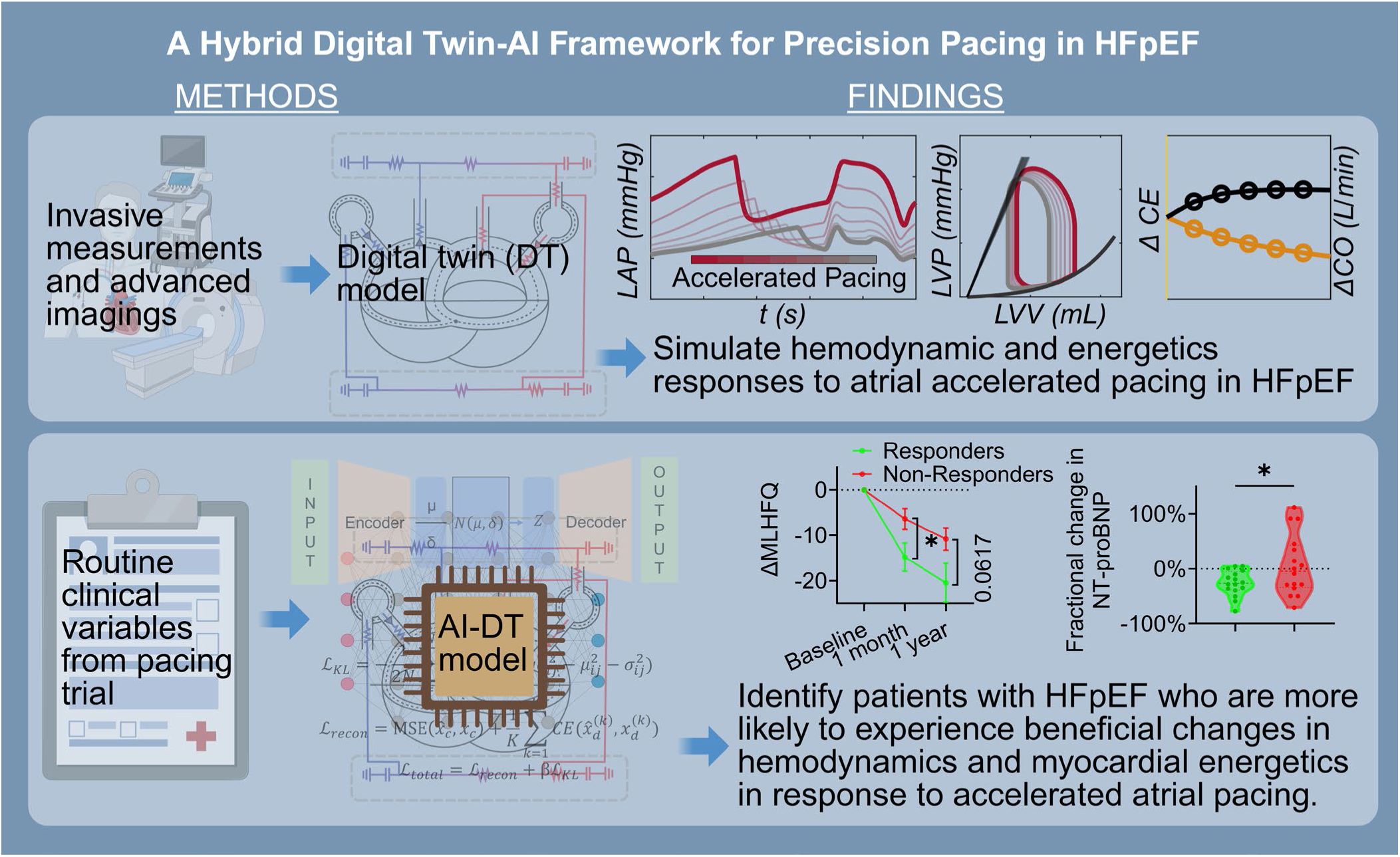

## Introduction

Heart failure with preserved ejection fraction (HFpEF) is a global public health crisis affecting millions worldwide, with rising costs, high hospitalization rates, diminished quality of life, and high mortality (1–3). Managing HFpEF remains a challenge, as most evidence-based therapies were developed for HFrEF (2), leaving patients with HFpEF with limited treatment options (4–7). While pharmacologic heart rate (HR) lowering benefits HFrEF (2, 8), β-blockers have shown neutral or adverse effects in HFpEF and are no longer recommended (2, 8–11). In addition, in patients with HFpEF and pre-existing pacemakers, the myPACE randomized trial showed that pacing at a moderately increased rate markedly improved quality of life, NT-proBNP levels, physical activity, and atrial fibrillation compared with standard settings (12). In contrast, the RAPID-HF randomized trial of rate-adaptive atrial pacing reported no improvement in exercise capacity or quality of life (13). Notably, both trials indicated a similar physiological effect, in which pacing shortened diastolic duration and led to a leftward and downward shift of the pressure–-volume loop, but offered divergent interpretations. myPACE attributed the resulting reduction in filling pressures to beneficial ventricular unloading at rest, whereas RAPID-HF considered the same response during physical activity detrimental because of impaired filling and reduced stroke volume (14, 15). While the seemingly conflicting results of these two trials are most likely due to the completely different interventions being studied, as well as differences in study design, endpoints, and patient populations (11,12), they also underscore the broader heterogeneity of HFpEF (16, 17). This heterogeneity is well-recognized across HFpEF therapies. For example, interatrial shunting exhibits clear responder and non-responder phenotypes that are strongly influenced by pulmonary vascular resistance and compliance (18). By extension, accelerated atrial pacing is also unlikely to exert uniform effects, and interpretation based on a single physiological marker may fail to account for the diverse mechanisms driving patient-specific responses.

Diastolic relaxation is an energy-consuming process requiring ATP for cross-bridge detachment and active calcium reuptake into the sarcoplasmic reticulum; thus, impaired energetic reserve can directly contribute to diastolic dysfunction (19, 20). Several studies have demonstrated that intrinsic mitochondrial abnormalities and compromised myocardial energetics with lower cardiac efficiency are common in patients with HFpEF (21–25) and these abnormalities are associated with worse outcomes (22, 26). Recent evidence further suggests that mitochondrial dysfunction and disrupted myocardial energy homeostasis may represent promising therapeutic targets in HFpEF (19, 23). Among available pharmacologic therapies, sodium-glucose cotransporter-2 (SGLT2) inhibitors show strong and consistent evidence for improving clinical outcomes in HFpEF (4, 5). Their benefits may involve, at least in part, improvements in myocardial energetics and cardiac efficiency (27), although the exact mechanisms remain under investigation. However, few studies have examined how pacing itself influences myocardial energetics. We therefore hypothesized that a favorable response to pacing therapy would be characterized by reduced myocardial oxygen demand and/or improved efficiency of left ventricular power output.

In this study, we developed a physiology-informed computational framework to identify HFpEF patients most likely to benefit from accelerated atrial pacing. Since invasive hemodynamic monitoring to assess individual pacing responses is clinically impractical, we employed an in silico approach using previously validated patient-specific cardiovascular modeling (digital twins) technology (28) to simulate physiological responses to pacing, including cardiac energy demand. These models predicted changes in key parameters: left atrial pressure (LAP), systolic blood pressure (SBP), cardiac output (CO), and cardiac efficiency (CE), defined as CO normalized to myocardial oxygen consumption. However, constructing digital twins requires comprehensive physiological data rarely available in clinical practice. To bridge this translational gap, we implemented a two-stage approach: first, we used generative AI trained on existing digital twins to generate a large virtual cohort; second, developing machine learning classifiers that predict pacing responses using only routine clinical variables. We hypothesized that patients with predicted improvements in cardiac efficiency, indicating favorable energetic adaptation to increased heart rate, would demonstrate superior clinical outcomes with pacing therapy. This hypothesis was then tested using the myPACE randomized trial, revealing that model-predicted physiological responses corresponded to improvements in quality of life, NT-proBNP levels, and functional capacity.

## Methods

### Study Population

This study utilized data from two complementary HFpEF cohorts, integrating real-world electronic health records (EHR) and randomized clinical trial datasets to develop and evaluate a physiology-informed digital twin–AI framework for predicting pacing response.

The first cohort, referred to as the HFpEF Digital Twin cohort (HFpEF-DT), was derived from a subset of patients included in our previous study of heart failure (28). The original cohort included patients across the full heart failure spectrum, and the present analysis focused on those with HFpEF (n = 146). This retrospective EHR-based cohort was drawn from the University of Michigan and the University of Wisconsin–-Madison. In that study, patient-specific digital twins were identified using EHR data, enabling detailed energetic and hemodynamic simulations. These data and corresponding digital twins were used to design the virtual pacing strategy and to train a physiology-informed generative AI (GenAI) model, implemented as a variational autoencoder (VAE), that learned the latent physiological representation of HFpEF patients and generated a large-scale virtual HFpEF cohort for AI model training.

The second cohort was obtained from a randomized clinical trial investigating pacing therapy in HFpEF: myPACE (NCT04721314) (12). The myPACE trial randomized patients with HFpEF and pre-existing pacemaker systems that minimized ventricular dyssynchrony to either a standard backup pacing rate of 60 bpm or an accelerated, personalized backup rate based on height and left ventricular ejection fraction. The primary endpoint was change in heart-failure–related quality of life, assessed by the Minnesota Living with Heart Failure Questionnaire (MLHFQ) at baseline, 1-month, and 1-year follow-up. Secondary endpoints included NT-proBNP levels, device-detected activity time as a measure of exercise capacity, and incidence of new-onset atrial fibrillation. Baseline clinical features (prior to any pacing intervention) were used to assess whether modeled physiologic response patterns corresponded to observed clinical outcomes. Specifically, only patients from the pacing arm of myPACE were included for validation (n = 48). Patients were categorized into simulated “response pattern” subgroups according to the digital-twin AI classifier output. Endpoints including quality-of-life scores, NT-proBNP levels, and device-detected activity were then used to assess the clinical relevance of these modeled physiological phenotypes.

### Digital Twin–Based Simulation of Accelerated Atrial Pacing

The digital twin model from our previous study was adapted to investigate the hemodynamic effects of accelerated atrial pacing. The digital twin model has been described in detail previously (28). In brief, the computational model includes both central and peripheral components. The central component, representing the heart, is based on the TriSeg model developed by Lumens et al. (29), which provides an idealized geometric framework for simulating passive and active myocardial mechanics, forming the basis for reproducing the impaired diastolic function characteristic of HFpEF. The peripheral component consists of lumped elements representing the atria, valves, pericardium, and systemic and pulmonary circulations. These components are coupled to the central TriSeg model to form a closed-loop cardiovascular system using an approach adapted from Kim et al. (30), enabling simulation of blood flow and pressure dynamics throughout the circulation. This previously established in silico environment allows precise parameter variation and has been shown to accurately reproduce heterogeneous (patho)physiological conditions in HFpEF (28, 31). Similar model configurations have been used for realistically simulating cardiac mechanics and hemodynamics under various pacing strategies (32, 33).

The virtual pacing strategy did not account for conduction-system abnormalities, which may partially explain inter-patient variability in therapeutic response of pacing (14). Atrial pacing was implemented in all digital-twin simulations, in simulations of both real patients and the virtual population. Ventricular activation followed atrial contraction after a fixed fraction of the cardiac cycle, representing the atrio-ventricular delay. The ratio between systolic and diastolic durations was varied with pacing rate according to an empirical relationship (34). The systemic venous compliance was adjusted across pacing rates to restore MAP toward baseline at steady state, approximating baroreflex-mediated regulation following changes in HR(35). Because myPACE lacked sufficient data for high-fidelity digital twin construction, pacing simulations were performed only in the HFpEF-DT and virtual cohorts. For each simulated patient, HR was increased from baseline to +30 bpm in increments of 5 bpm.

This framework enabled simulation of detailed hemodynamic and myocardial oxygen demand changes under accelerated atrial pacing. We focused on four primary variables: LAP, SBP, CO, and CE. LAP was chosen as a surrogate of left-ventricular filling pressure which is reflective of diastolic function; SBP captured contractile performance, which is increasingly recognized as impaired in some HFpEF phenotypes (16); and CO was included because RAPID-HF demonstrated that pacing can shorten filling time and reduce output. However, these parameters often lead to opposing interpretations of the same physiological phenomenon: a shorter diastolic duration can be viewed as beneficial unloading when focusing on LAP, but as a detrimental loss of stroke volume when focusing on CO (36). It should be noted, however, that during exercise the centralization of blood volume and changes in body position (e.g., recumbent vs upright) complicate this scenario, such that the net effect on LAP and CO may differ from resting conditions and require careful interpretation. To integrate these apparently conflicting perspectives, we evaluated CE, which is defined as CO divided by left ventricular myocardial oxygen demand (LVMVO₂), as a comprehensive indicator of how effectively the heart converts metabolic energy into forward flow. LVMVO₂ was estimated from the LV pressure–volume area (PVA) following the formulation of Suga et al. (37) and Takaoka et al. (38). We hypothesized that favorable response to pacing would be characterized by improved CE, reflecting reduced energetic cost for maintaining output, whereas decreased CE would indicate a maladaptive response. The detailed virtual pacing protocol and quantification of PVA are described in the Supplement, and the source code, developed in MATLAB (R2024b, MathWorks Inc.), is available on GitHub: https://github.com/beards-lab/SimulatePacing.git.

### The AI Model Development and Validation

The virtual pacing strategy described above was applied to patients from the HFpEF-DT cohort and to the virtual cohort, both of which contained fully established digital twins. As these patients had not undergone pacing therapy, our goal was to develop a framework that could map simulated pacing responses onto routinely available clinical variables and predict responses in real pacing trial, linking simulated changes in LAP, SBP, CO, and CE with observed clinical benefit. Since the myPACE trial contained insufficient data to enable full digital twin construction or direct virtual pacing simulation, we sought to enable an AI model to learn the behavior of the digital twin. Initially, classifiers were trained using the HFpEF-DT cohort to map baseline clinical characteristics restricted to variables also available in the myPACE trial to simulated pacing-induced changes in LAP, SBP, CO, and CE. However, the limited sample size (146 patients) was insufficient for robust model training. To address this limitation, an expanded virtual HFpEF population was generated using a Gen AI and used in place of the HFpEF-DT cohort to train classifiers mapping baseline clinical features to pacing-induced hemodynamic changes. The overall strategy is illustrated in Figure 1 using CE prediction as an example.

**Figure 1.**
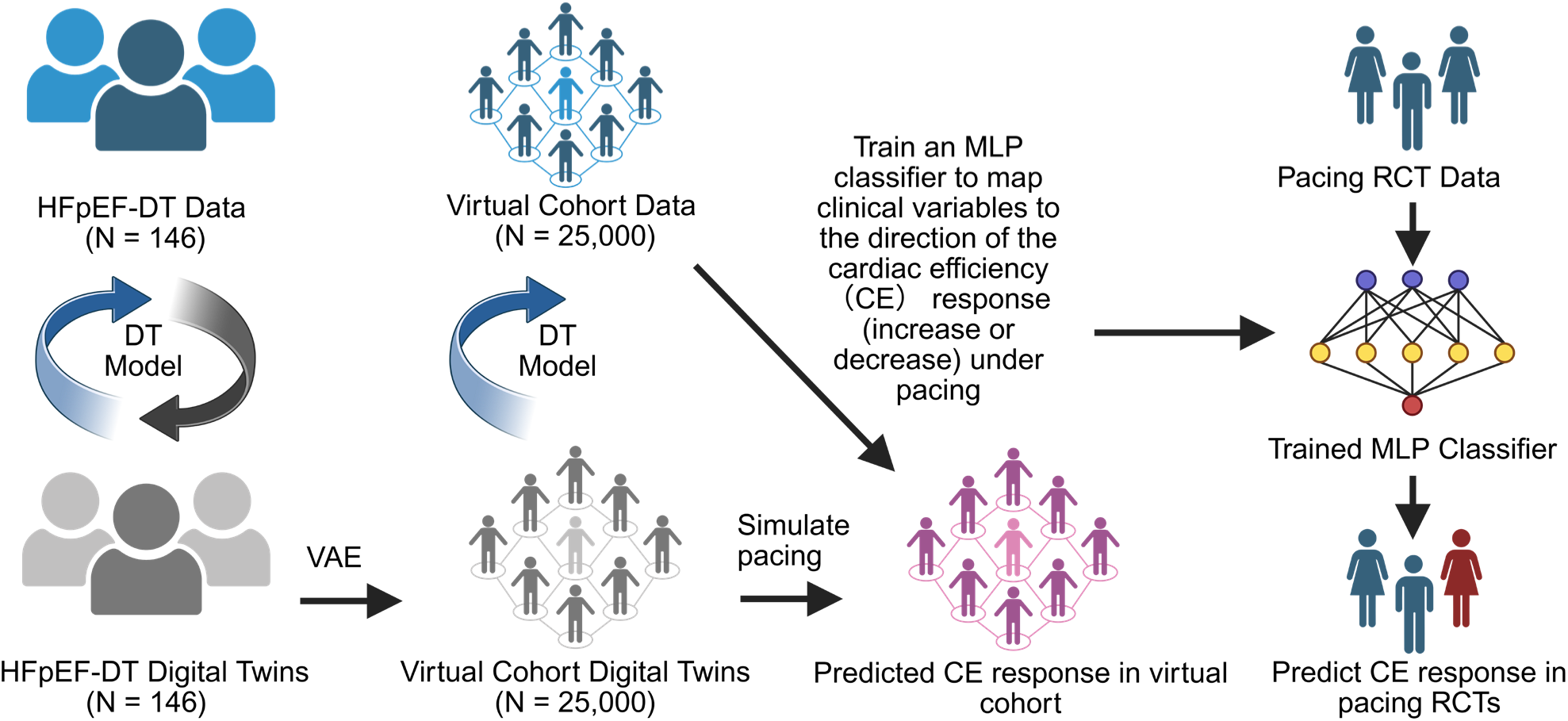
Proposed workflow integrating a physiology-informed digital twin (DT) model and artificial intelligence (AI) to predict pacing-induced changes in cardiac efficiency (CE) in patients with HFpEF. Patient-specific digital twins were derived from the HFpEF-DT cohort (N = 146) and used to train a variational autoencoder (VAE), which generated a virtual cohort’s digital twins (N = 25,000). The DT model was then used to simulate the corresponding virtual clinical data and pacing responses in this virtual population to derive CE-based response labels. A multilayer perceptron (MLP) classifier was subsequently trained to map baseline clinical variables to the direction of CE change (increase or decrease). The trained classifier was then applied to the myPACE randomized controlled trial (RCT) dataset to predict physiological responses and identify likely responders.

We chose a VAE (39) as the Gen AI model, trained on digital twins from the HFpEF-DT cohort. This design allows the VAE to learn the latent physiological representation of HFpEF patients. The trained VAE was then used to generate digital twins forming a virtual HFpEF cohort, on which the digital twin model was applied to simulate corresponding hemodynamics at baseline and in response to increased pacing rate. The resulting virtual cohort served as the training set for a fully connected multilayer perception (MLP) with two hidden layers and a sigmoid output unit. The MLP input included the subset of virtual cohort variables that overlapped with routinely available clinical measurements in the myPACE trial, comprising demographic information, vital signs, and standard echocardiographic parameters (17 variables in total). The corresponding digital twins provided simulated pacing responses in LAP, SBP, CO, and CE, which were used to derive binary labels: LAP, CO, and CE were labeled based on whether they increased or decreased, while SBP was labeled according to whether its reduction exceeded 8.5 mmHg. This cutoff was chosen to balance the training dataset (∼50% responders vs. non-responders), rather than to define a clinically meaningful threshold.

These four labels served as classification outputs, each trained with the same input features but using independent classifiers. The HFpEF-DT cohort was used as the testing set to evaluate classifier performance in predicting simulated changes in LAP, SBP, CO, and CE. The HFpEF-DT and virtual cohorts contained no missing values. For the myPACE trial, missing data were imputed using the k-nearest neighbors (KNN) algorithm with k = 5. The VAE and MLP classifiers were implemented in Python (v3.11.8) using the PyTorch framework. Model performance was evaluated using multiple metrics from scikit-learn library. Principal component analysis of the real and VAE-generated cohorts, along with calculation of the mean pairwise Euclidean distance, was used to characterize cohort-level diversity and assess whether the VAE showed any evidence of mode collapse. Details of the model architecture and training procedures are provided in the Supplementary Data, and the source code is available on GitHub: https://github.com/beards-lab/PhysicalAI.git.

### Statistical Analysis

All statistical analyses, except for the linear mixed model, were performed using GraphPad Prism version 10.2.2 (GraphPad Software, LLC). Continuous variables are presented as mean ± SD or SEM. Between-group comparisons were conducted using unpaired *t*-tests. Repeated-measures data were analyzed with mixed-effects models using restricted maximum likelihood (REML) estimation, with time, simulated response pattern group, and their interaction specified as fixed effects and subjects as random effects. When significant main or interaction effects were detected, Tukey’s multiple-comparison test was applied for post hoc analysis. Linear mixed models were implemented in MATLAB R2025b (MathWorks Inc.) using the statistics and machine learning toolbox. Two-sided *P* values < 0.05 were considered statistically significant (\**p* < 0.05, \*\**p* < 0.01, \*\*\**p* < 0.001, and \*\*\*\**p* < 0.0001).

For NT-proBNP, because the change from baseline to follow-up visits was not normally distributed, percentage change from baseline was analyzed instead. Patients with NT-proBNP values within the normal range at both baseline and follow-up were excluded from analyses involving NT-proBNP but retained for analyses of other endpoints (e.g., quality-of-life or functional measures). This analytic approach is consistent with the original randomized trial and avoids overinterpretation of minimal biomarker variability.

## Results

### Physiology-Informed Digital Twins Predict Heterogeneous Hemodynamic and Energetic Responses to Accelerated Atrial Pacing in HFpEF

Digital twins of patients from the HFpEF-DT cohort were previously identified in our earlier study (28). Using these individualized models, we simulated hemodynamic and energetic responses to accelerated atrial pacing.

Figure 2 illustrates virtual pacing responses from a canonical healthy subject (Panel A, as defined in our previous work) and from two representative patients with HFpEF from the HFpEF-DT cohort (patient A in Panel B and patient B in Panel C). Under baseline sinus rhythm, the patient digital twins reproduced the key pathophysiological feature of HFpEF, impaired diastolic function, manifested as elevated LAP. During virtual pacing, with HR increased stepwise up to +30 bpm from baseline, the healthy subject’s average LAP changed minimally (6.25 vs 5.72 mmHg), whereas both patient A and B exhibited substantial reductions (18.55 vs 12.09 mmHg and 25.82 vs 17.21 mmHg, respectively), consistent with invasive hemodynamic observations in HFpEF (14, 40–45). Simulated pressure-volume (PV) relationships demonstrated leftward shifts of PV loops along the end-systolic and end-diastolic pressure-volume relations (ESPVR and EDPVR) in all subjects.

**Figure 2.**
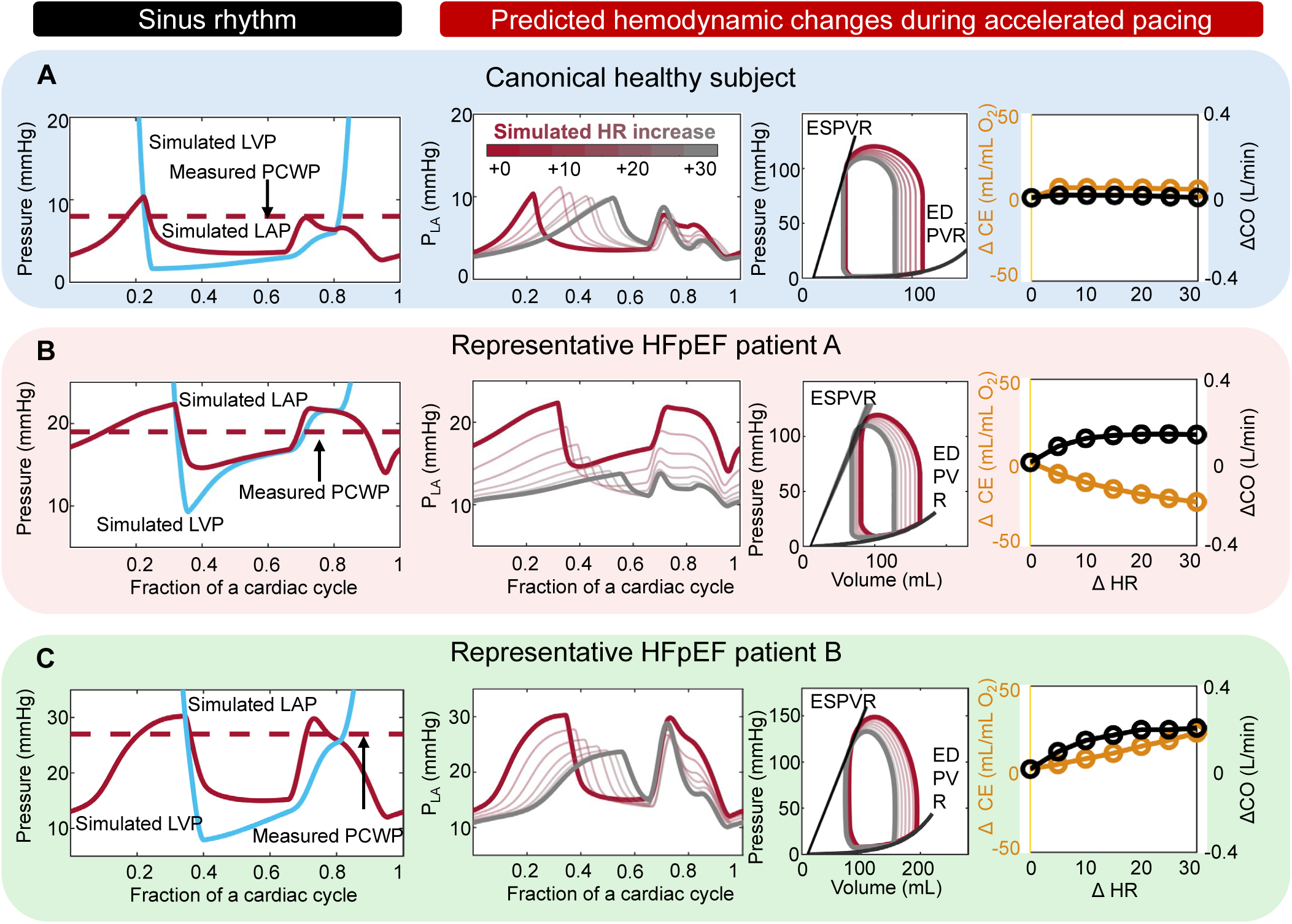
Predicted hemodynamic and energetic responses to accelerated atrial pacing in a canonical healthy subject and two representative patients with HFpEF. Each panel (row) corresponds to one subject: (A) a canonical healthy subject, (B) representative HFpEF patient A, and (C) representative HFpEF patient B. From left to right within each panel: (1) simulated left ventricular pressure (LVP, blue) and left atrial pressure (LAP, red) under sinus rhythm, with measured pulmonary capillary wedge pressure (PCWP, dashed line) shown for reference; (2) simulated LAP waveforms during stepwise heart-rate increases from baseline to +30 bpm (color transition from red to gray); (3) left-ventricular pressure–volume (PV) loops at corresponding pacing rates, showing end-systolic (ESPVR) and end-diastolic (EDPVR) relations; and (4) predicted changes in cardiac output (ΔCO, black, right axis) and cardiac efficiency (ΔCE, yellow, left axis) as functions of pacing rate (ΔHR, bpm).

Because HFpEF is characterized by a disproportionally steep EDPVR, the modeled filling pressure decreased more prominently under pacing, accompanied by a modest reduction in stroke volume, which is also consistent with prior acute hemodynamic measurements (13, 40, 43, 44). The simulations further showed a decline in SBP and pulse pressure (PP) during pacing in both healthy and HFpEF models. Patient A exhibited a magnitude of SBP and PP reduction (116.39 vs 107.32 mmHg; 53.39 vs 39.77 mmHg) similar to that of the healthy subject (116.93 vs 107.30 mmHg; 43.85 vs 29.42 mmHg), whereas patient B showed a slightly greater decline (SBP 120.81 vs 109.74 mmHg; PP 64.16 vs 47.55 mmHg; Supplementary Figure 1). CO remained stable in the healthy subject but increased modestly in patient A (∼0.15 L/min) and patient B (∼0.20 L/min) despite the reduced stroke volume.

Energetic analysis revealed that LVMVO₂ remained nearly unchanged in the healthy model but increased in patient A and decreased in patient B (Supplementary Figure 1), leading to a reduction in CE (CE = CO/LVMVO₂) for patient A and an increase for patient B. This indicates that although pacing modestly improves predicted CO in both patients, their predicted energetic responses diverge. Specifically, the HFpEF-DT model predicts that patient A requires a disproportionately greater increase in LVMVO₂ to maintain cardiac output, reflecting a less energy-efficient adaptation, whereas patients like patient B achieved similar or even improved output with reduced or only minimally increased oxygen consumption, suggesting a more favorable energetic response to pacing.

Each of the 146 HFpEF-DT patients was virtually paced using the digital twin model. Among them, 141 patients (96.6%) exhibited a decrease in LAP during accelerated atrial pacing at one or more pacing rates (Figure 3A, green lines), whereas 5 patients (3.4%) showed paradoxical LAP elevation (Figure 3A, red lines). All patients showed a reduction in SBP, though the magnitude varied: 79 patients (54%) showed a drop ≥ 8.5 mmHg (Figure 3B, green lines), while 67 (46%) demonstrated smaller reductions (Figure 3B, red lines). CO responses were also heterogeneous: 137 patients (94%) exhibited an increase in CO at one or more pacing rates compared with baseline (Figure 3C, green lines), whereas 9 (6%) showed a continuous decline throughout pacing (Figure 3C, red lines). Finally, 97 patients (66%) showed decreased CE with pacing (Figure 3D, red lines), while 49 (34%) demonstrated improved CE (Figure 3D, green lines).

**Figure 3.**
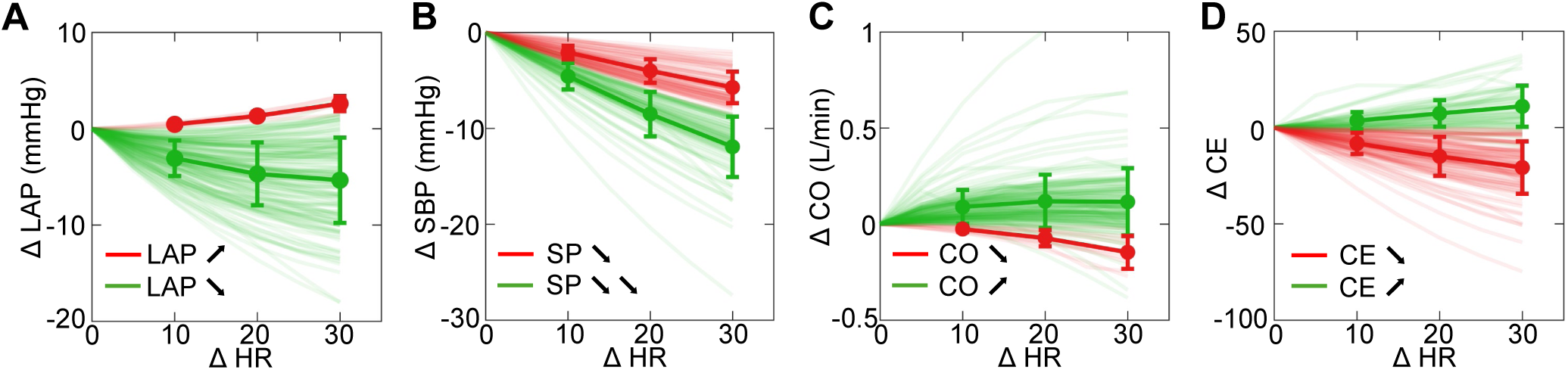
Heterogeneous hemodynamic and energetic responses to accelerated atrial pacing across the HFpEF-DT cohort. Each line represents the simulated response of one patient’s digital twin as heart rate (ΔHR) is increased from baseline to +30 bpm. Colors indicate favorable versus unfavorable response directions for each physiological endpoint rather than simple increases or decreases. For ΔLAP (A), increases are shown in red and decreases in green. For ΔSBP (B), substantial systolic pressure drops (ΔSBP > −8.5 mmHg) are shown in green, whereas smaller drops or increases are shown in red. For ΔCO (C) and ΔCE (D), increases are shown in green and decreases in red. Bold lines and error bars represent the cohort mean ± SD.

Together, these simulations highlight substantial hemodynamic and energetic heterogeneity across HFpEF patients, motivating a framework to relate these modeled patterns to clinical outcomes.

### Physiology-Informed AI Partially Learned the Digital Twin Model Behavior

The results above demonstrate that accelerated atrial pacing elicited markedly different hemodynamic and energetic responses among patients with HFpEF. To evaluate whether these simulated responses could help explain why patients benefit from pacing or not, we next examined data from patients who received accelerated atrial pacing therapy in the myPACE trial. Because patients in these trials lacked sufficient physiological data to construct high-fidelity digital twins directly, we applied a physiology-informed AI–digital twin framework to predict pacing-induced hemodynamic and energetic changes in these real-world cohorts.

A VAE (Supplementary Figure 2) was trained using digital twins from the HFpEF-DT cohort to generate a large virtual population (n = 25,000). The generated digital twins exhibited similar probability distributions to those of the HFpEF-DT cohort (Supplementary Figure 3), and pairwise correlations between physiological parameters were well preserved (Supplementary Figure 4). Using the digital twin model, each virtual twin was used to generate corresponding simulated clinical data. These virtual data displayed a wider but smoother distribution compared with the original HFpEF-DT data, while maintaining comparable physiological ranges (Supplementary Figure 5). Principal component analysis was performed on data from both the HFpEF-DT cohort and the VAE-generated virtual population. The synthetic cohort occupied a similar region of the physiological feature space as the real patients, with comparable overall spread, indicating that the generative model preserved physiological heterogeneity (Supplementary Figure 6). After normalization, the mean pairwise Euclidean distances of the real and virtual cohorts were 5.76 and 5.73, respectively, demonstrating that the VAE maintained cohort-level diversity and did not exhibit mode collapse. The pacing simulations of the virtual cohort showed overall consistent hemodynamic and energetic responses with the HFpEF-DT cohort, although with larger magnitudes of change (Supplementary Figure 7). Specifically, 95.6% of virtual patients exhibited a decrease in LAP under accelerated atrial pacing, 93.8% showed an increase in CO, 36.1% demonstrated improved CE, and 47.0% had an SBP reduction greater than 8.5 mmHg.

These categorical changes (increase/decrease or large/small drop) in LAP, SBP, CO, and CE were then used as binary outcome labels to train four independent multilayer perceptron MLP classifiers.

Since all classifiers used the same input features, correlations between labels were examined. As shown in Supplementary Figure 8, the four labels were moderately correlated, indicating that individual patients often exhibited concordant directional changes (e.g., all increasing or all decreasing) across these parameters, meaning that the four classification tasks captured largely overlapping physiological response patterns.

The virtual cohort was used as the training set, and the HFpEF-DT cohort served as the testing set to evaluate classifier performance in predicting digital twin–derived hemodynamic and energetic responses to accelerated atrial pacing. Model performance was assessed using standard metrics, including accuracy, precision, recall, F1 score, and the area under the receiver operating characteristic (ROC) curve (AUC). ROC curves for the four classifiers are shown in Figure 4, and detailed performance metrics are summarized in Table 1. Although the AUC values for predicting changes in LAP (0.97 vs 0.74) and CO (0.91 vs 0.70) declined when moving from training to testing data, both classifiers maintained high overall accuracy (LAP: 0.96 vs 0.97; CO: 0.93 vs 0.90) and precision (LAP: 0.99 vs 0.97; CO: 0.97 vs 0.95). The apparent reduction in AUC likely reflects the extreme class imbalance in the testing set (>90% positive cases) and the small number of negative samples, which can make ROC-based metrics unstable despite stable predictive performance. These results indicate that the MLP classifiers were able to learn physiologically meaningful patterns from the digital twin simulations and generalized well to unseen HFpEF patients.

**Figure 4.**
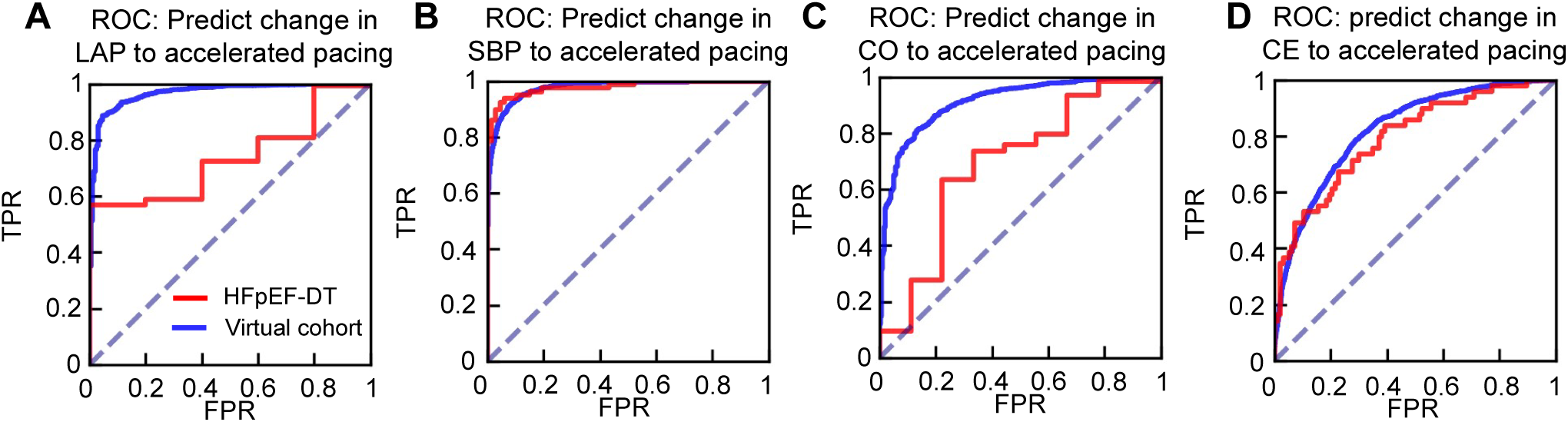
AI Model performance in predicting digital twin–derived hemodynamic and energetic responses to accelerated atrial pacing. Receiver operating characteristic (ROC) curves for classifiers predicting changes in (A) left atrial pressure (LAP), (B) systolic blood pressure (SBP), (C) cardiac output (CO), and (D) cardiac efficiency (CE) in response to accelerated atrial pacing. Blue curves represent model performance in the virtual training cohort (n = 25,000), and red curves represent performance in the HFpEF-DT testing cohort (n = 146). The classifiers achieved high discriminative performance in training and maintained acceptable generalization to the testing cohort despite class imbalance.

**Table 1:** Performance metrics of MLP classifiers in predicting pacing-induced hemodynamic and energetic changes. Each classifier (LAP, SBP, CO, CE) predicts the direction of pacing-induced change in the corresponding variable. Values are shown for the training (outside parentheses) and testing (in parentheses) datasets. LAP, left atrial pressure; SBP, systolic blood pressure; CO, cardiac output; CE, cardiac efficiency; AUC, area under the receiver operating characteristic curve.

| Classifiers | AUC | Accuracy | Precision | Recall | F1 |
| --- | --- | --- | --- | --- | --- |
| LAP | 0.97 (0.74) | 0.96 (0.96) | 0.99 (0.97) | 0.97 (0.99) | 0.98 (0.98) |
| SBP | 0.91 (0.98) | 0.95 (0.88) | 0.84 (0.98) | 0.89 (0.78) | 0.89 (0.87) |
| CO | 0.91 (0.70) | 0.93 (0.90) | 0.97 (0.95) | 0.96 (0.94) | 0.96 (0.95) |
| CE | 0.82 (0.79) | 0.75 (0.73) | 0.71(0.59) | 0.54 (0.61) | 0.62 (0.6) |

### Digital Twin–AI Framework Distinguishes Pacing Responders from Non-Responders in myPACE

The trained classifiers were applied to predict pacing-induced hemodynamic and energetic responses in patients from the myPACE trial.

In the myPACE trial, 40 of 48 patients with HFpEF were predicted to have decreased LAP under pacing, 23 were predicted to exhibit a SBP reduction greater than the cutoff, 42 were predicted to have increased CO, and 17 were predicted to have increased CE. We next examined whether these predicted physiological responses were associated with clinical outcomes in the myPACE cohort. As shown in Figure 5, patients predicted to have favorable hemodynamic or energetic changes (decreased LAP, larger SBP drop, increased CO, or increased CE) tended to demonstrate greater improvements in quality of life (MLHFQ score) at both 1-month and 1-year follow-up. Among these, only the groups predicted to have a larger SBP reduction or increased CE showed significantly greater MLHFQ improvement at 1-month follow-up compared with their counterparts (Figure 5B, 5D). We also assessed the relationship between predicted responses and changes in NT-proBNP at 1 month. Only patients predicted to have a larger SBP decrease exhibited significantly greater reductions in NT-proBNP (Figure 5F), whereas other predicted labels showed no significant association (Figure 5E, 5G, 5H). No significant differences were observed in pacemaker-detected physical activity between any predicted subgroups (Supplementary Figure 9).

**Figure 5.**
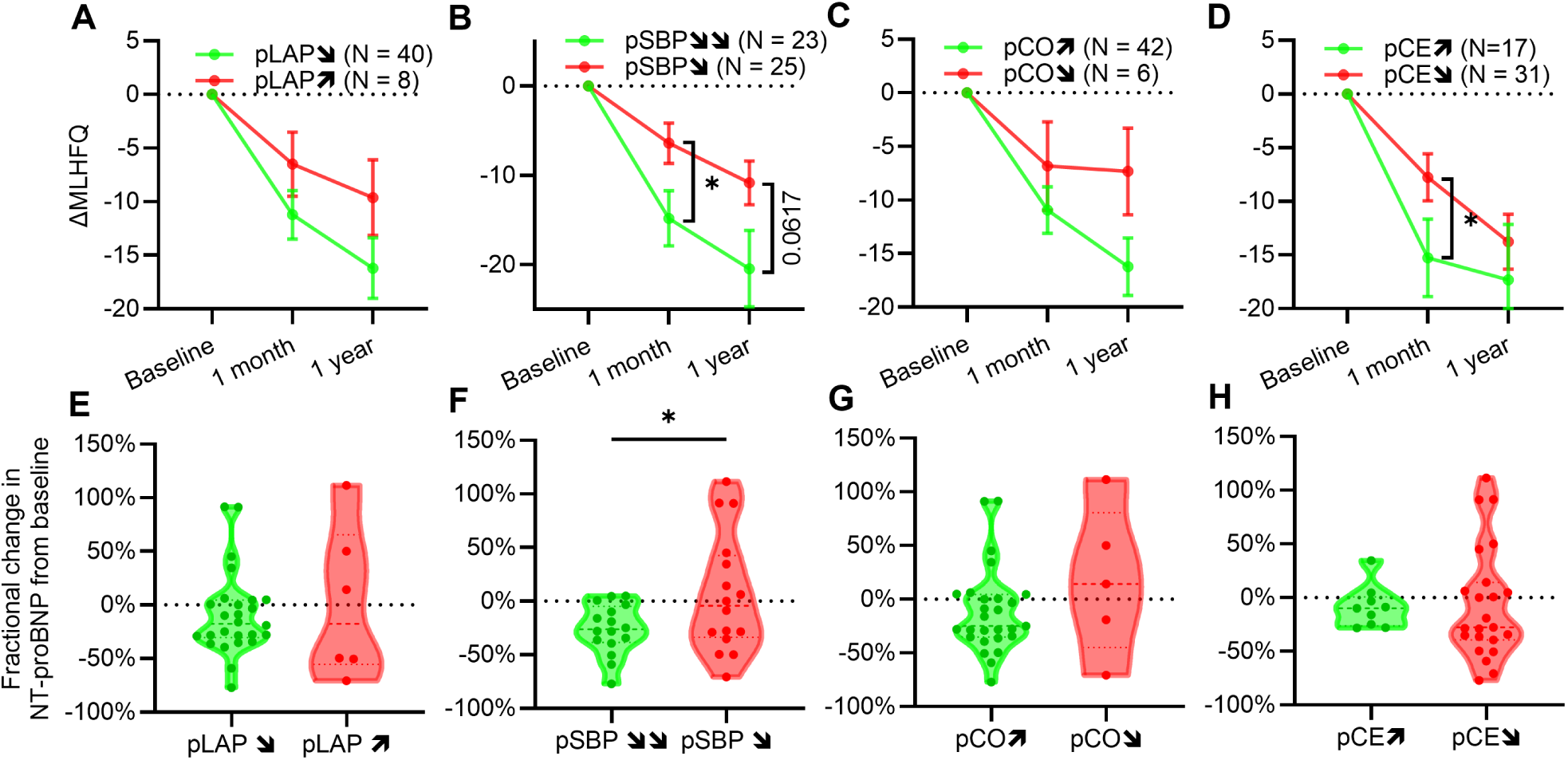
Associations between AI-predicted pacing responses and functional and biomarker outcomes in the myPACE trial. Panels A–D show changes in quality-of-life score (ΔMLHFQ) at 1-month and 1-year follow-up, stratified by AI-predicted hemodynamic and energetic responses to accelerated atrial pacing: (A) predicted change in left atrial pressure (pLAP), (B) systolic blood pressure (pSBP), (C) cardiac output (pCO), and (D) cardiac efficiency (pCE). Patients predicted to have a larger drop in SBP or increased CE showed significantly greater MLHFQ improvement at 1-month. Panels E–H show the corresponding percentage change in NT-proBNP at 1 month, grouped by predicted LAP (E), SBP (F), CO (G), and CE (H) responses. A larger predicted SBP reduction was associated with a greater decrease in NT-proBNP (*p < 0.05*). Data are shown as mean ± SEM (A–D) and as violin plots (E–F).

Given the class imbalance in model outputs, which could weaken statistical power in the above analysis, we further evaluated the continuous output probabilities of each classifier. These probabilities quantify the likelihood that a patient belongs to the favorable physiological-response class (e.g., CE and CO improvement or larger SBP reduction). They were analyzed using linear mixed-effects models for longitudinal endpoints (MLHFQ and pacemaker-detected activity) and simple linear regression for changes in NT-proBNP. Consistent with the previous analysis, a higher probability of a larger SBP decrease or increased CE was associated with greater MLHFQ improvement at 1-month follow-up. In addition, a higher predicted probability of a larger SBP drop was significantly correlated with a greater reduction in NT-proBNP (Supplementary Table 1). Moreover, the probability of increased CE correlated with greater improvements in pacemaker-detected physical activity at 6 months. Collectively, results from the myPACE trial indicate that patients predicted to have increased CE experienced the most consistent improvements in both symptoms and activity levels, and those predicted to have larger SBP reductions showed greater symptoms and biomarker improvements. These findings support an association between modeled physiologic response patterns and observed clinical benefit, but they do not imply prospective predictive capability.

## Discussion

In this translational study, we integrated physiology-based digital twin modeling with state-of-the-art AI approaches to identify HFpEF patients most likely to benefit from accelerated atrial pacing. By leveraging simulated hemodynamic and energetic responses from the digital twin framework, the model identified simulated physiological signatures that corresponded to clinical signals observed in the myPACE trial, using only routinely available clinical variables.

Our findings challenge a “one-size-fits-all” perspective that views HR modulation as uniformly beneficial or detrimental in HFpEF, no matter through pharmacologic reduction or pacing acceleration. Instead, the results highlight the physiological heterogeneity underlying pacing responses. Notably, improvement in CE emerged as a key discriminator of modeled therapeutic benefit, identifying patients who exhibited greater symptom relief, larger reductions in NT-proBNP, and increased pacemaker-detected physical activity (Figure 5, supplementary figure 9). Patients with pacing-induced CE improvement maintained or augmented cardiac output without a disproportionate rise in myocardial oxygen demand, as reflected by a concurrent fall in SBP and PP (Figure 3, supplementary figure 7).

These findings suggest that improved cardiac efficiency may represent a unifying physiological marker of favorable pacing response, bridging energetic and hemodynamic perspectives and providing a mechanistically interpretable biomarker for patient selection. Moreover, the integration of AI with physiology-based digital twins enables robust, generalizable modeling using limited and easily obtainable clinical data, thereby enhancing the translational potential of this framework for future applications.

### Heart Rate Modulation and Therapeutic Strategy in HFpEF

Heart-rate modulation has long been viewed as a cornerstone of heart failure management. In HFrEF, β-blockers and ivabradine improve outcomes by reducing myocardial oxygen consumption, prolonging diastolic filling time, enhancing coronary perfusion, and reversing maladaptive remodeling (2, 8).

However, these physiological benefits do not translate to HFpEF, where diastolic stiffness rather than impaired systolic contractility predominates (46–48). In a stiff ventricle, prolonging diastole increases filling pressure and wall stress rather than stroke volume, as reflected by the steeper EDPVR (Figure 2) (46). Consequently, clinical trials and secondary-analyses have failed to demonstrate benefit from β-blockers in HFpEF (9, 10, 49), and some even reported higher rates of adverse cardiovascular events, hospitalization, and new-onset atrial fibrillation (11, 50). These findings support recent guideline recommendations that routine β-blocker use for rate control in HFpEF should be avoided in the absence of another clear indication (2).

An alternative strategy, heart-rate acceleration, has been explored in several recent studies (51–53). Approximately 20% of patients with HFpEF have implanted pacemakers for bradyarrhythmia, providing an opportunity to test device-based heart-rate modulation (54). The myPACE trial demonstrated that moderate atrial pacing improved quality of life, reduced NT-proBNP, increased physical activity, and lowered burden of atrial fibrillation and may lower major cardiovascular adverse events in patients with pre-existing pacemakers (12, 55). In contrast, the RAPID-HF trial found no improvement in exercise capacity or quality of life, likely because rate-adaptive pacing failed to increase cardiac output in patients with impaired stroke volume under pacing (13). Both studies, however, revealed similar underlying hemodynamic patterns, which is shortened diastolic duration and a leftward/downward shift of the pressure-volume loop. but they offered opposing interpretations of whether this represents beneficial unloading or detrimental underfilling.

Taken together, these findings indicate that neither heart-rate reduction nor universal pacing acceleration constitutes a one-size-fits-all strategy in HFpEF. The optimal direction of heart-rate modulation likely depends on patient-specific hemodynamic reserve and energetic efficiency, which are features represented mechanistically in our framework. Although this study focuses on accelerated atrial pacing in HFpEF, our results also provide hypothesis-generating insight into the opposite end of the spectrum, namely heart-rate reduction. As our strategy classifies HFpEF patients based on their simulated, patient-specific physiological responses to heart-rate elevation, it may help identify individuals who are likely to benefit from β-blocker therapy. Indeed, among 146 patients in the HFpEF-DT cohort, 45 had paroxysmal atrial fibrillation and may benefit from β-blocker therapy. Of these, only 12 were classified as having CE improvement with accelerated atrial pacing. This initial finding suggests that individualized assessment of hemodynamic and energetic profiles may better guide heart-rate modulation strategies. While the fidelity of our mechanistic simulations is relatively crude at this stage, the approach may eventually help frame a more individualized approach to inform personalized decision-making modulation or a more refined patient selection strategy for future trials.

### Divergent Hemodynamic and Energetic Responses to Accelerated Atrial Pacing

Direct measurement of elevated filling pressure remains the gold standard for diagnosing HFpEF, and alleviating elevated filling pressure is an overarching goal in HFpEF management. Therefore, assessing how pacing affects filling pressure provides valuable mechanistic insight. In our simulations, more than 95% of patients in the HFpEF-DT cohort exhibited a decrease in LAP during accelerated atrial pacing (Figure 3A, Supplementary Figure 7), while the remaining patients showed only a minor (<2 mmHg) increase. These findings are consistent with invasive hemodynamic studies in humans, which demonstrated a modest but reproducible reduction in LAP during pacing in most patients with HFpEF (14, 40–45).

One hallmark of HFpEF pathophysiology is elevated filling pressure due to passive ventricular stiffness arising from altered cardiomyocyte and extracellular matrix properties, which steepen the EDPVR (Figure 1). Pacing reduces LAP not by modifying this intrinsic stiffness, but by shortening diastolic filling time, effectively shifting the pressure–volume loop along the existing abnormal EDPVR. Although this maneuver lowers LAP, it simultaneously reduces stroke volume—a finding reproduced in our simulations and observed in prior hemodynamic studies (13, 40, 43, 44), despite CO was maintained or slightly increased in most simulated HFpEF patients (Figure 3C, Supplementary Figure 7), consistent with some invasive measurements (40, 44). This suggests that pacing achieves hemodynamic unloading primarily through temporal redistribution rather than structural improvement. In other words, pacing lowers filling pressure by abbreviating diastole, not by improving compliance.

However, as the pacing rate continues to rise, diastolic duration becomes insufficient for adequate ventricular filling, leading to a progressive reduction in stroke volume and, ultimately, a reduced cardiac output. Under these conditions, the myocardium may exhibit oxygen consumption that is unchanged or even increased, resulting in a decline in cardiac efficiency. This pattern is consistent with our simulation results (Figure 3D) and may explain why rate-adaptive pacing increases peak HR without improving exercise capacity (13).

Unlike the hemodynamic effects of pacing, the energetic consequences of heart-rate modulation remain poorly defined in patients with HFpEF. In vitro studies using failing and mechanically stiffer cardiomyocytes have shown that higher pacing rates markedly increase oxygen consumption, not only through enhanced calcium cycling and cross-bridge activation but also because diastolic relaxation itself becomes an additional, energetically costly process (56). However, very few in vivo studies have investigated how LVMVO₂ changes with atrial pacing, particularly in HFpEF. Earlier cardiac resynchronization therapy studies demonstrated that total LVMVO₂ increases after resynchronization, while the rise in external work was often proportionally greater, resulting in net gains in mechanical efficiency (57–59). Subsequent analyses, however, revealed substantial heterogeneity: in non-responders, LVMVO₂ frequently increased without corresponding improvement in work output, yielding neutral or even negative changes in efficiency (60). Animal experiments have also shown that increased diastolic wall tension can elevate oxygen consumption and reduce overall efficiency (61), as relaxation itself becomes an additional energy-consuming process.

Our study reveals a similar phenomenon in HFpEF during atrial pacing. The overall energetic demand increased, but the degree of change in CE varied markedly among individuals. This heterogeneity represents, to our knowledge, the first systematic prediction of divergent energetic responses to pacing in HFpEF. Although direct LVMVO₂ measurements were not available, the AI-digital twin framework provided a physiologically interpretable surrogate: changes in simulated CE effectively distinguished patients who derived greater symptomatic and biochemical benefit in randomized trials from those who did not (Figure 5).

### Energetic Demand and Metabolic Efficiency as a Therapeutic Target in HFpEF

Metabolic dysregulation is increasingly recognized as a central feature of HFpEF. Phenomapping studies consistently identify a subgroup of HFpEF patients characterized by obesity, insulin resistance, and other metabolic abnormalities, underscoring the tight link between systemic metabolism and myocardial performance (17, 62). At the myocardial level, multiple studies have demonstrated impaired energetic efficiency and reduced cardiac energy reserve in HFpEF (21–25). Experimental work suggests that these abnormalities arise from decreased mitochondrial oxidative capacity, reduced glucose oxidation, and a compensatory increased myocardial glycolysis rate (24). Diastolic relaxation itself is an energy-demanding process, requiring ATP for cross-bridge detachment and calcium reuptake into the sarcoplasmic reticulum. Consequently, impaired energetic reserve can directly exacerbate diastolic dysfunction (19, 20). Beyond diastole, subtle impairments in systolic contractile reserve are also increasingly appreciated as contributors to HFpEF pathophysiology (16), reflecting the broader energetic inefficiency of the myocardium.

Among current pharmacologic treatments, SGLT2 inhibitors provide a consistent benefit in HFpEF (4, 5). Although the precise mechanisms remain incompletely understood, accumulating evidence suggests that SGLT2 inhibitors enhance myocardial energetic efficiency largely through systemic metabolic effects, including a shift toward greater glucose and ketone oxidation and yielding more ATP per unit of oxygen consumed (27). Notably, because SGLT2 expression in human myocardium is minimal, these energetic benefits are likely mediated indirectly rather than through direct myocardial SGLT2 inhibition. Clinically, HFpEF patients with metabolic comorbidities appear to derive the greatest benefit from SGLT2 inhibition (63). Similarly, preclinical studies have shown that glucagon-like peptide-1 (GLP-1) receptor agonists can improve myocardial energetics and efficiency in experimental models (64).

These findings collectively reinforce the notion that impaired myocardial energy utilization and reduced efficiency are not merely epiphenomena but potential therapeutic targets in HFpEF. Within this context, our observation that pacing-induced improvements in cardiac efficiency identify responders suggests that restoring energetic balance, whether through metabolic or electrophysiologic modulation may represent a convergent pathway toward therapeutic benefit in HFpEF.

### Clinical Implications and Future Directions

From a technical perspective, our unique framework bridges the complementary strengths of physiology-based digital twins and data-driven AI while mitigating their inherent limitations. Digital twin models offer mechanistic interpretability and physiological insight but often rely on high-dimensional data, idealized assumptions, and computationally expensive simulations. In contrast, data-driven approaches are mathematically robust and computationally efficient once trained, but they typically lack interpretability and demand large training populations. The present framework bridges the two paradigms: a generative AI expands the virtual sample space, while a classifier learns physiological patterns from the digital twin, enabling efficient, mechanistically grounded mapping of simulated responses onto clinically available variables using only limited and easily obtained clinical data. This approach thus overcomes the typical trade-off between mechanistic fidelity and practical deployability. Future work will focus on deeper physical-AI integration, where the governing equations of the digital twin are directly embedded as physics-informed constraints within neural networks (65). Such a design would retain the full physiological knowledge base of the digital twin while dramatically accelerating computation and enabling continuous learning from clinical data.

From a clinical standpoint, the current model provides a mechanistic framework rather than a predictive tool, offering a pragmatic way to explore patient stratification in device-based HR modulation therapy. Among all simulated hemodynamic and energetic variables, improvement in CE emerged as the physiologically meaningful discriminator of pacing response. Notably, a greater SBP reduction, a simple, readily measurable clinical parameter, paralleled increases in CE, reflecting the same underlying mechanism whereby the heart maintains cardiac output with reduced energetic demand.

This finding suggests that pacing-induced SBP change may serve as an easily obtainable surrogate of CE improvement, linking model-derived physiology to bedside applicability in an exploratory manner.

Beyond pacing, the framework may extend to broader domains of HR modulation, including pharmacologic HR control, although not all mechanisms are symmetric. Unlike pacing, β-blockers affect not only HR but also myocardial metabolism, remodeling, and autonomic tone. Understanding how these pathways interact with hemodynamic and energetic phenotypes of HFpEF represents a natural next step. Future studies should also investigate how pacing influences myocardial metabolism *in vivo*, as the energetic consequences of HR modulation remain incompletely characterized. Together, these directions outline a translational roadmap for combining digital physiology and AI to achieve precision pacing and, more broadly, personalized HR modulation in HFpEF.

This study provides a unique physiology-informed framework and was able to identify patients who demonstrated greater functional improvement in the myPACE trial. However, several considerations and limitations should be noted. First, the clinical evaluation was retrospective and restricted to patients in the pacing arm of myPACE. Therefore, the findings reflect association. The AI model is not trained on patient-level outcomes but instead predicts physiological responses to pacing therapy in individual patients. Since the key hemodynamic and energetic responses were not measured, future studies incorporating more comprehensive physiological measurements including simultaneous hemodynamic, metabolic, and device-derived signals will be important to refine and validate the findings. Second, although the classifiers were trained on a large virtual HFpEF population, the underlying physiological structure originates from a relatively small cohort (n = 146), which may limit the diversity of HFpEF phenotypes represented in the virtual cohort. Future work using larger HFpEF datasets together with simple phenotype-based augmentation strategies may help broaden the diversity of the virtual population. Finally, the number of myPACE patients with complete baseline and follow-up data is modest, reducing statistical power and limiting the ability to evaluate interactions among simulated physiological variables. Larger prospective studies, ideally integrating pacing protocols with detailed physiological profiling, will be needed to determine whether simulated phenotypes correspond to true physiological subgroups and to test whether this framework can ultimately support clinical decision-making for heart-rate modulation in HFpEF.

## Conclusion

The current study demonstrates that integrating physiology-based digital twin modeling with AI enables mechanistic exploration of hemodynamic and energetic responses to accelerated atrial pacing in HFpEF. Improvement in simulated cardiac efficiency emerged as a mechanistically grounded and hypothesis-generating, rather than clinically validated, marker of favorable pacing response, with corresponding reductions in systolic blood pressure providing an easily measurable clinical correlate of these modeled patterns. These findings support the concept that precision pacing, which is guided by patient-specific energetic and hemodynamic profiles, may represent a new therapeutic avenue for HFpEF. Future prospective studies integrating this framework into pacing trials will be essential to determine whether these modeled physiologic signatures correspond to true biological subgroups and whether they can ultimately contribute to personalized, physiology-informed device therapy for patients with HFpEF.

## Supporting information

Supplemental Detailed Methods

## Acknowledgements

We thank Dr. Farhan Raza (Department of Medicine–Cardiovascular Disease, University of Wisconsin–Madison) for his contributions to the clinical care and original data collection of patients included in the previously published HFpEF-DT cohort used in this study. We also thank Andrew J. Meyer, whose earlier work on digital-twin model generation contributed to a portion of the dataset incorporated from the prior publication.

## Ethical Approval

Ethical approval for the HFpEF-DT cohort was obtained from the Institutional Review Boards of the University of Michigan (IRB protocol number HUM00243399) and the University of Wisconsin–Madison (IRB protocol number 2019-0535). The MyPACE cohort received approval from the Institutional Review Board of the University of Vermont Medical Center.

## Data Availability

Non–PHI data from the HFpEF-DT cohort have been made publicly available in our previous publication(28). The virtual cohort generated by the GenAI model is available in the corresponding Git repository (https://github.com/beards-lab/PhysicalAI.git). Data from the myPACE randomized clinical trial is available from the original corresponding investigators upon reasonable request.

## Notes

### Competing Interest Statement

The authors have declared no competing interest.

### Clinical Trial

NCT04721314

### Funding Statement

This study was funded by NIH grant HL173346 and the Michigan Medicine-PKUHSC Joint Institute for Translational and Clinical Research. We also acknowledge support from the Michigan Biology of Cardiovascular Aging Postdoctoral Fellowship, which provided funding for the training and career development of Feng Gu during this study.

### Author Declarations

IRB committees of the University of Michigan, the University of Wisconsin-Madison, and the University of Vermont gave ethical approval for this work.

