## Supplemental Detailed Methods for "Physiology-Informed Digital Twin-AI Framework Predicts Pacing Therapy Response in HFpEF"

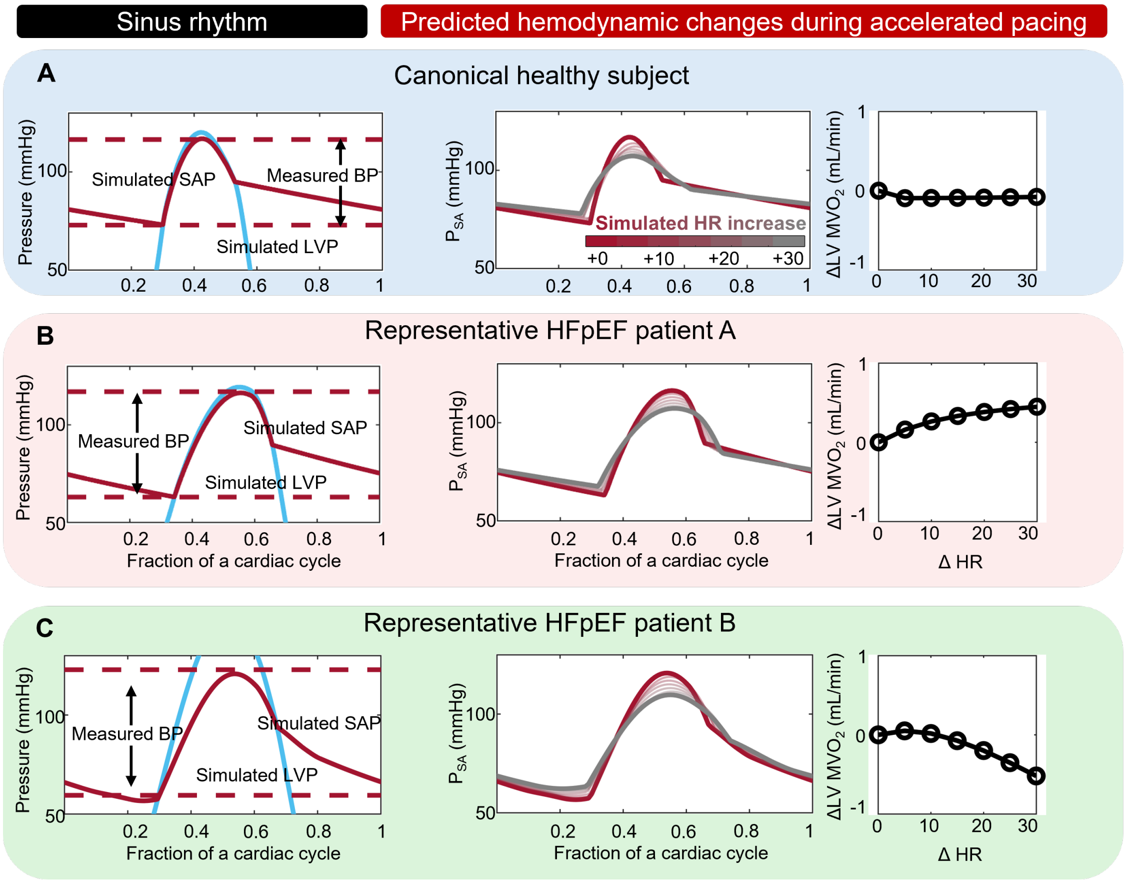

**Supplementary Figure 1: Predictive changes in systolic blood pressure and myocardial oxygen consumption during accelerated atrial pacing in a canonical healthy subject and two representative HFpEF patients.** Each panel corresponds to one subject: (A) a canonical healthy subject, (B) representative HFpEF patient A, and (C) representative HFpEF patient B. From left to right within each panel: (1) simulated systolic arterial pressure ($SAP$, red) and left-ventricular pressure ($LVP$, blue) during sinus rhythm, with measured brachial systolic blood pressure (dashed line) shown for reference; (2) simulated $SAP$ waveforms during stepwise heart-rate increases from baseline to $+30$ bpm (color transition from red to gray); (3) predicted changes in left-ventricular myocardial oxygen consumption ($\Delta \mathrm{LVMVO_2}$) as a function of pacing rate ($\Delta HR$, bpm).

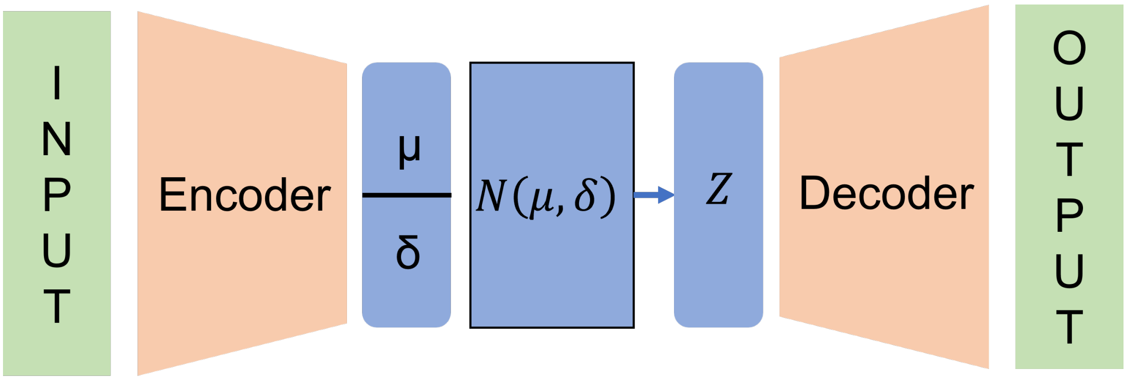

**Supplementary Figure 2: Schematic structure of the variational autoencoder (VAE)**

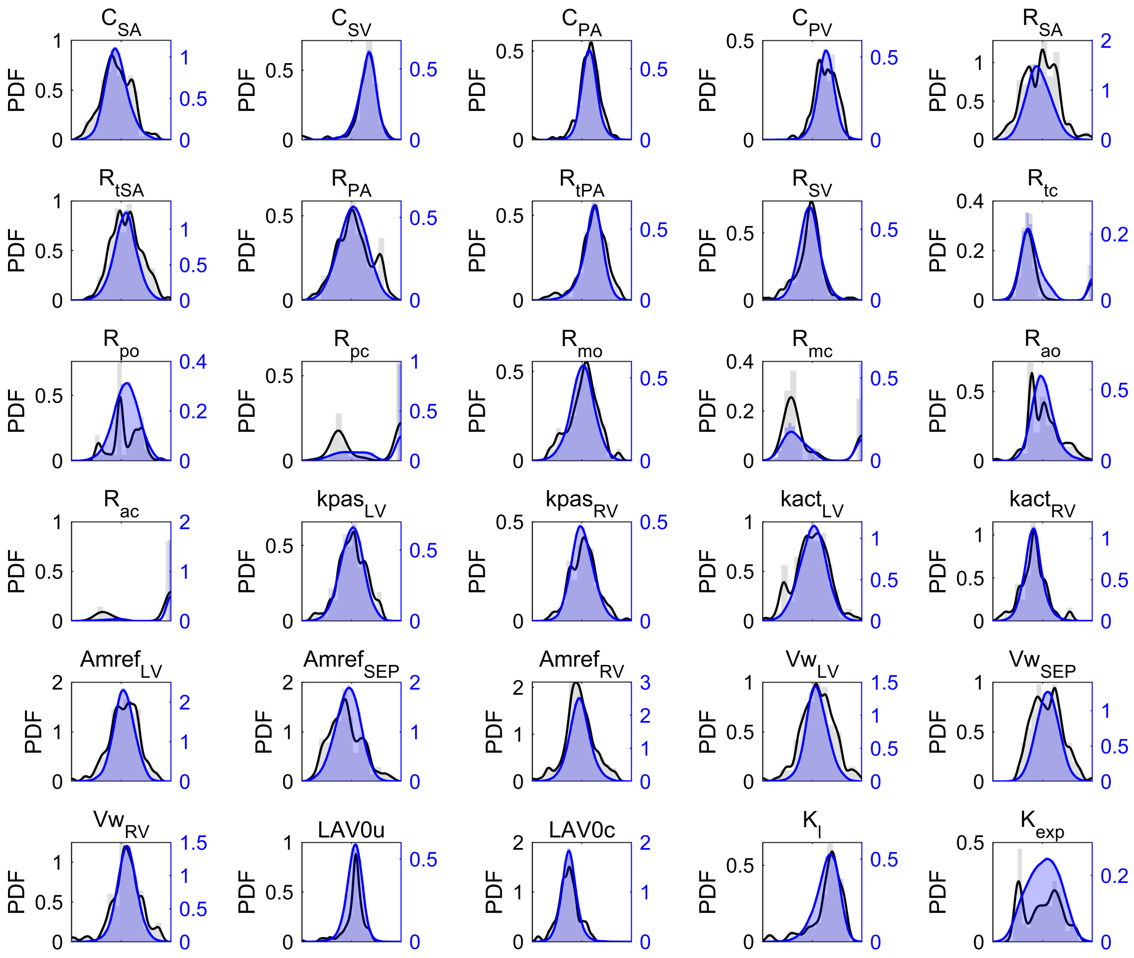

**Supplementary Figure 3: Distributional comparison of model parameters between real HFpEF digital twins and Gen-AI–generated virtual cohort digital twins.** Each subplot represents one digital-twin model parameter across 146 HFpEF patients from the HFpEF-DT cohort, showing the empirical distribution of log-transformed real digital-twin parameter values (black; left y-axis) and the corresponding distribution of log-transformed Gen-AI–generated parameter samples (blue; right y-axis). For each parameter, shaded histograms display probability-density estimates, and solid lines depict kernel-smoothed probability density functions (PDFs), allowing visual assessment of agreement between real and generated digital-twin parameter distributions.

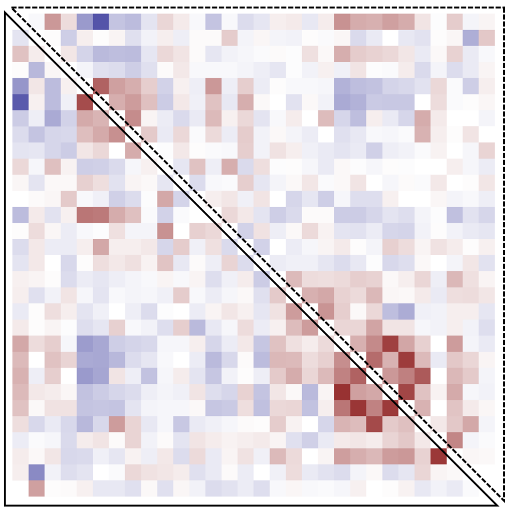

**Supplementary Figure 4: Comparison of correlation matrices between real HFpEF digital twins and Gen-AI–generated virtual-cohort digital twins** The lower-left triangle shows pairwise correlations among calibrated digital twins of 146 HFpEF patients from HFpEF-DT, whereas the upper-right triangle displays correlations from the 25,000-patient virtual cohort generated by the variational autoencoder. Colormap intensity reflects the magnitude and direction of the correlation coefficient (red indicates stronger positive correlation; blue indicates stronger negative correlation), enabling visual comparison of correlation structure between real and generated digital twins.

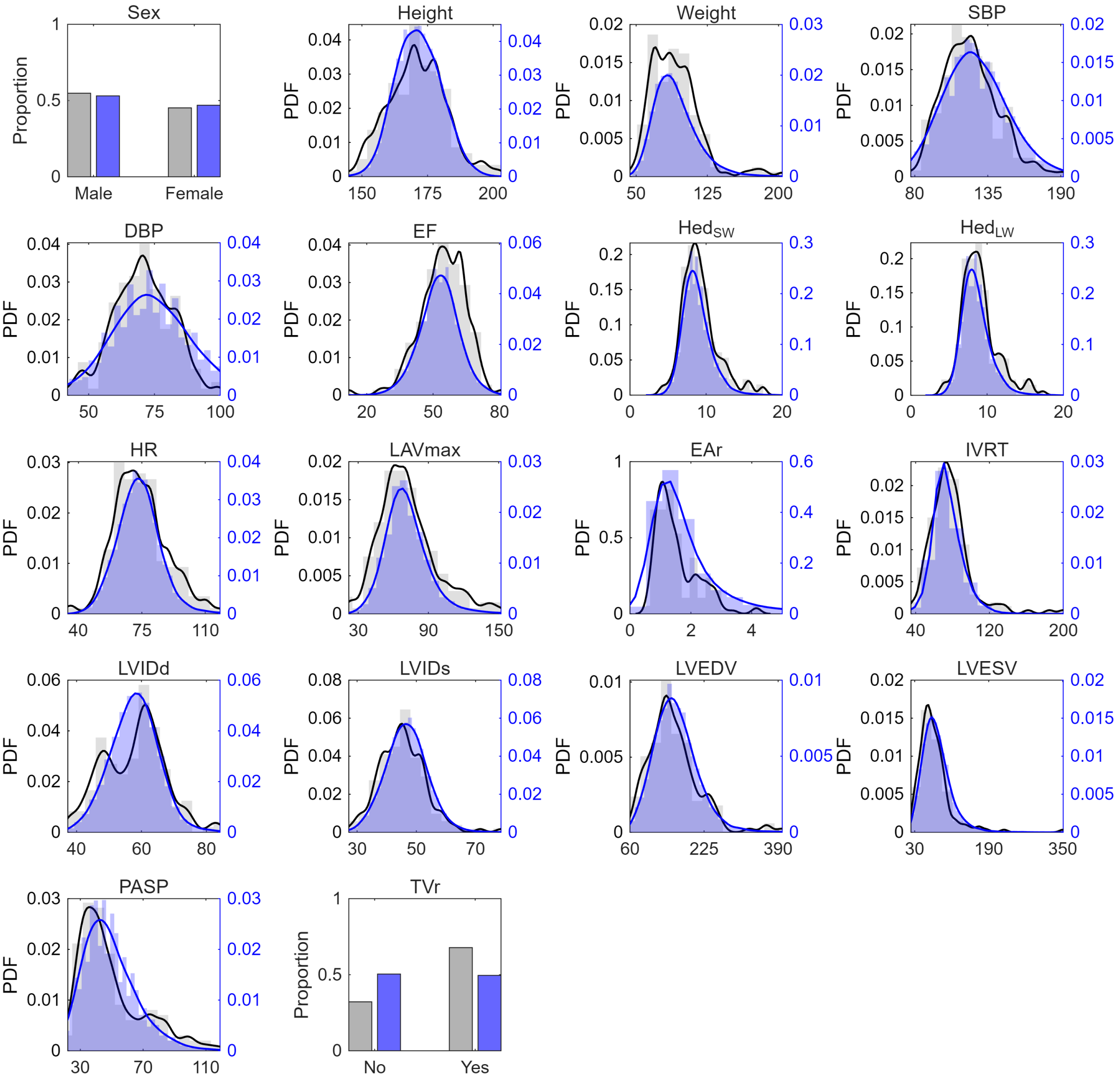

**Supplementary Figure 5: Comparison of real clinical measurements from the HFpEF-DT cohort and digital-twin–derived virtual measurements obtained by propagating Gen-AI–generated virtual-cohort digital twins through the digital twin model.** Each subplot represents one routinely collected demographic, vital-sign, or echocardiographic variable, showing the empirical distribution from real HFpEF-DT patients (black; left y-axis) and the corresponding distribution of DT-generated virtual clinical data (blue; right y-axis). Continuous variables are displayed using histogram-based and kernel-smoothed probability density functions, whereas categorical variables—Sex and TVr (presence of tricuspid regurgitation)—are shown as proportions. This analysis evaluates how well the physiologically informed virtual data recapitulate the marginal distributions observed in the HFpEF-DT cohort.

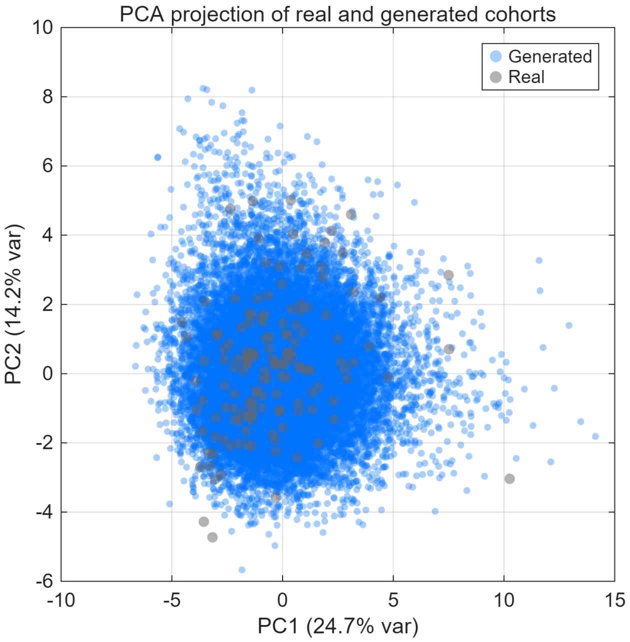

**Supplementary Figure 6: PCA visualization of real and virtual clinical data.** Principal component analysis (PCA) of the combined dataset, showing the distribution of real HFpEF-DT patients (gray) and digital twin–generated virtual patients (blue) projected onto the first two principal components. Each point represents an individual patient in the reduced-dimensional feature space. The spread and arrangement of points reflect the underlying multivariate structure of the clinical variables used to construct the virtual cohort.

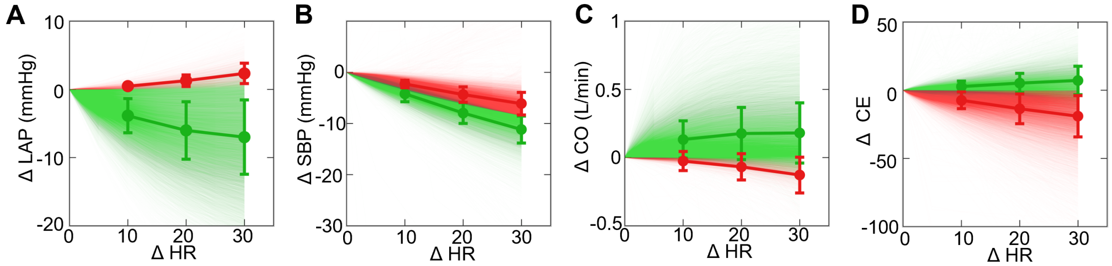

**Supplementary Figure 7: Heterogeneous hemodynamic and energetic responses to accelerated atrial pacing across the virtual cohort.** Each line represents the simulated response of one virtual digital twin as heart rate ($\Delta HR$) is increased from baseline to $+30\,\mathrm{bpm}$. Colors indicate favorable versus unfavorable response directions for each endpoint rather than simple increases or decreases. For $\Delta\mathrm{LAP}$ (A), increases are shown in red and decreases in green. For $\Delta\mathrm{SBP}$ (B), substantial systolic pressure drops ($\Delta\mathrm{SBP} > -8.5\,\mathrm{mmHg}$) are shown in green, whereas smaller drops or increases are shown in red. For $\Delta\mathrm{CO}$ (C) and $\Delta\mathrm{CE}$ (D), increases are shown in green and decreases in red. Bold lines and error bars indicate the cohort mean $\pm$ SD.

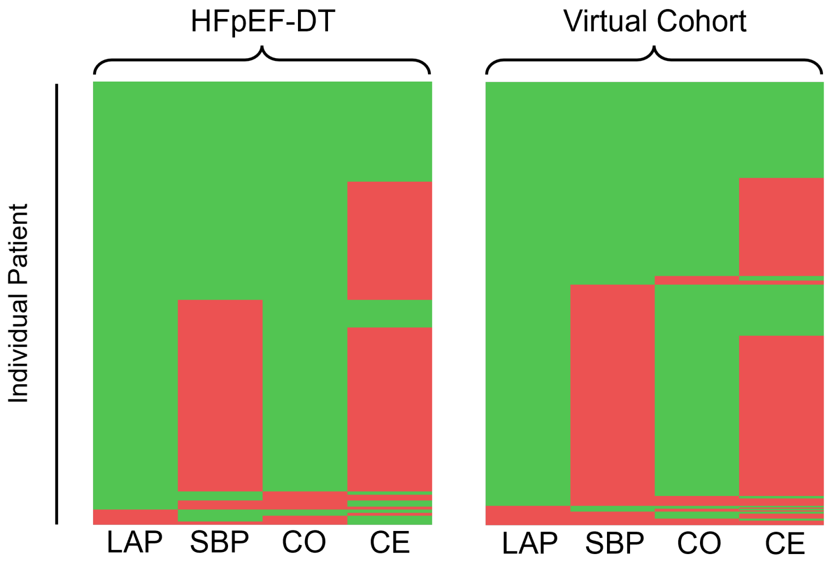

**Supplementary Figure 8: Individual-level consistency of pacing-response direction across hemodynamic and energetic responses in the HFpEF-DT cohort and the virtual cohort.** Each row represents a single patient (HFpEF-DT, left) or virtual patient (virtual cohort, right), and each column corresponds to one pacing-response metric: $\Delta\mathrm{LAP}$, $\Delta\mathrm{SBP}$, $\Delta\mathrm{CO}$, and $\Delta\mathrm{CE}$. Colors follow the same physiologic response-direction conventions used in Figure 3 and Supplementary Figure 6: $\Delta\mathrm{LAP}$ increases are shown in red and decreases in green; substantial systolic pressure drops ($\Delta\mathrm{SBP} > -8.5, \mathrm{mmHg}$) are shown in green, whereas smaller drops or increases are shown in red; and for both $\Delta\mathrm{CO}$ and $\Delta\mathrm{CE}$, increases are green and decreases are red. This visualization illustrates whether individual patients demonstrate consistent favorable or unfavorable pacing-response patterns across multiple physiologic measures.

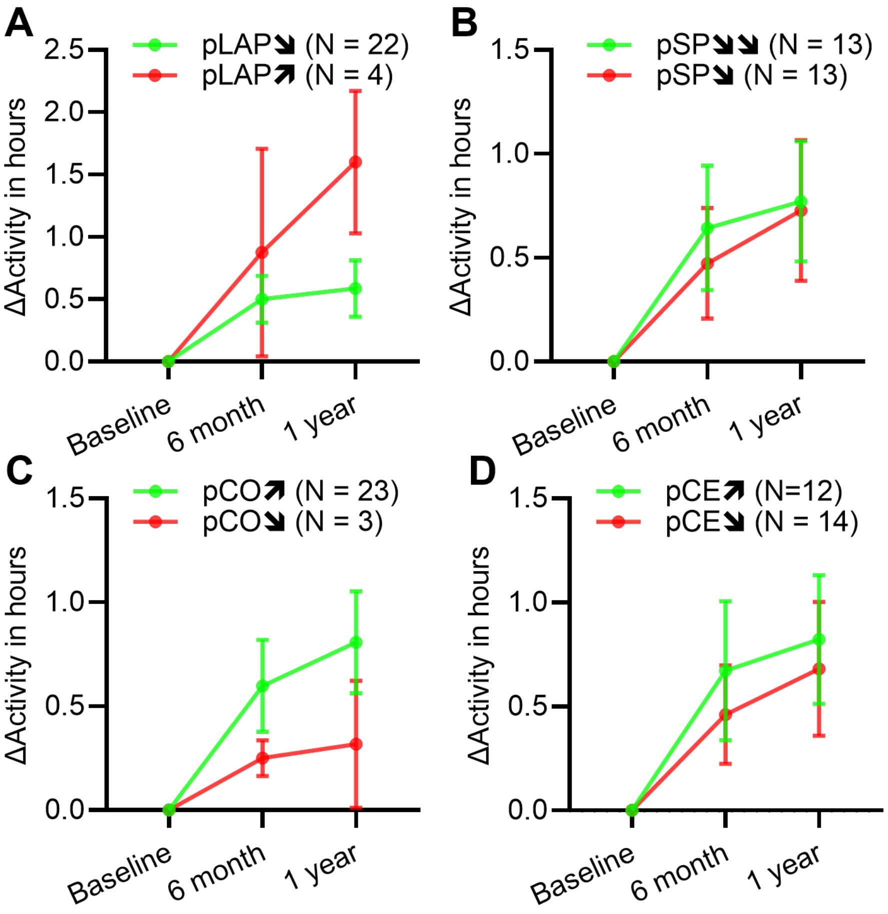

**Supplementary Figure 9: Associations between AI-predicted pacing responses and functional and biomarker outcomes in the myPACE trial.** Panels A–D show changes in pacer-detected daily activity (Δactivity) at 6-month and 1-year follow-up, stratified by AI-predicted hemodynamic and energetic responses to accelerated atrial pacing: (A) predicted change in left atrial pressure ($p_{\mathrm{LAP}}$), (B) systolic blood pressure ($p_{\mathrm{SBP}}$), (C) cardiac output ($p_{\mathrm{CO}}$), and (D) cardiac efficiency ($p_{\mathrm{CE}}$).

**Supplementary Table 1**

#### Associations between classifier-predicted probability and pacing-induced changes in clinical endpoints

| Classifiers | MLHFQ$_{1m}$ | MLHFQ$_{1y}$ | NT-proBNP | Activity$_{6m}$ | Activity$_{1y}$ |
|---|---|---|---|---|---|
| LAP | -13.14 (0.2761) | -14.27 (0.2381) | 0.13 (0.492) | 0.91 (0.4343) | -2.28 (0.0295)* |
| SBP | -8.12 (0.043)* | -7.67 (0.0771) | -0.38 (0.0307)* | 0.45 (0.1834) | 0.22 (0.4938) |
| CO | 5.09 (0.7837) | -3.67 (0.8465) | 0.07 (0.7111) | 1.04 (0.6179) | -0.89 (0.6891) |
| CE | -12.00 (0.0363)* | -5.35 (0.3858) | -0.17 (0.35) | 1.08 (0.0430)* | 0.28 (0.6147) |

For MLHFQ and pacer-detected activity, continuous classifier probabilities were evaluated using linear mixed-effects models to account for repeated measurements over time. Values shown are the estimated fixed-effect coefficients ($\beta$), with $p$-values in parentheses. NT-proBNP was assessed only at the 1-month follow-up; therefore, associations were quantified using Pearson correlation, reported as correlation coefficients ($r$) with $p$-values in parentheses. Statistical significance is indicated by an asterisk (*).

### Supplementary method 1: Details of Digital Twin–Based Simulation of Accelerated atrial pacing

#### Brief description of the digital twin model

The digital twin model used in this study was adapted from our previous work [1]. Briefly, it is a biophysical, mechanistic model that derives chamber pressure and flow from sarcomere-level tension generation, integrated with a simplified three-dimensional cardiac geometry and a lumped-parameter (0D) circulation model. This formulation provides an optimal balance between computational efficiency and physiological fidelity, allowing the model to reproduce key clinical characteristics measured by advanced imaging and invasive hemodynamics.The model is implemented in MATLAB and is publicly available at: `https://github.com/beards-lab/TriSeg-Digital-Twins.git`.

Compared with the previous version, the major update in the current implementation is the revised pericardial pressure formulation. In the earlier model, pericardial pressure was described by:

$$P_{\text{peri}} = \exp\left(\frac{V_{\text{total}}}{V_{0,\text{total}}} K_{\text{PC}}\right) + B_{\text{PC}}, \tag{1}$$

where $P_{\text{peri}}$ denotes the pericardial pressure, $V_{\text{total}}$ denotes the total cardiac volume, and $V_{0,\text{total}}$ represents the maximal total cardiac volume for each individual, computed as the sum of ventricular and atrial chamber volumes at end-diastole plus the corresponding wall volumes.

In the updated model used in this study, the pericardium is modeled as:

$$P_{\text{peri}} = K_1 \exp\left[K_2 \left(\frac{V_{\text{total}}}{V_{0,\text{total}}} - 1\right)\right], \tag{2}$$

which provides a smoother and more physiologically consistent dependence of pericardial pressure on total cardiac volume, improving numerical stability during pacing simulations.

#### Strategy for Patient-Specific Simulation of Accelerated Atrial Pacing

The digital twin model is used to predict patient-specific hemodynamic and energetic responses to accelerated atrial pacing. A key physiological assumption is that, following an abrupt increase in heart rate, the cardiovascular system rapidly attains a new quasi-steady hemodynamic equilibrium through autonomic nervous system–mediated regulation. Consistent with acute invasive studies [2–8], this shift is characterized by relatively stable arterial pressure, reduced cardiac filling pressures, and modest changes in cardiac output.

To reproduce this behavior, the digital twin model adjusts the compliance of the systemic veins, defined as

$$C_{\text{SV}} = \frac{V_{\text{SV}}}{P_{\text{SV}}}, \tag{3}$$

where $V_{\text{SV}}$ is the stressed venous volume and $P_{\text{SV}}$ is the venous pressure. Reducing $C_{\text{SV}}$ shifts blood volume toward the venous compartment, decreasing ventricular filling and thus lowering filling pressures while preserving mean arterial pressure (MAP) during pacing.

Because sarcomere tension development in the model is entirely driven by the prescribed activation function, accelerated pacing requires adjusting the timing of this function to reflect the physiological observation that systole and diastole both shorten with increased heart rate, but diastole shortens more. We retain the baseline piecewise activation form, governed by the fractions $k_{TS}$ and $k_{TR}$ of the cardiac cycle:

$$Y(t) = \begin{cases} 0.5\left(1 - \cos\left(\frac{\pi t}{k_{TS}}\right)\right), & 0 \le t \le k_{TS}, \\ 0.5\left(1 + \cos\left(\frac{\pi (t - k_{TS})}{k_{TR}}\right)\right), & k_{TS} \le t \le k_{TS} + k_{TR}, \\ 0, & \text{otherwise.} \end{cases} \tag{4}$$

At baseline, we set $k_{TS} = 0.35$ and $k_{TR} = 0.15$ based on physiological data [9]. To convert these fractions into absolute timing at any heart rate (HR), we use empirical expressions for the QS2 interval (electromechanical systole) [10]:

$$\mathrm{QS2} = \begin{cases} 545.17 - 2.117\,\mathrm{HR}, & \text{male}, \\ 546.5 - 2.0\,\mathrm{HR}, & \text{female}. \end{cases} \tag{5}$$

and model the isovolumic relaxation time as

$$\mathrm{IVRT} = 70 \times \frac{75}{\mathrm{HR}}. \tag{6}$$

The total duration of myocardial active tension is then

$$\mathrm{ActT} = \mathrm{QS2} + \mathrm{IVRT}, \tag{7}$$

which represents the full activation interval rather than the purely mechanical systolic period. Assuming a 2:1 ratio between systolic and diastolic activation durations, the model assigns

$$k_{TS} = \frac{2}{3}\left(\frac{\mathrm{ActT}\cdot\mathrm{HR}}{60000}\right), \qquad k_{TR} = \frac{1}{3}\left(\frac{\mathrm{ActT}\cdot\mathrm{HR}}{60000}\right). \tag{8}$$

These combined adjustments—modulation of systemic venous compliance and heart-rate–dependent activation timing—allow the digital twin to reproduce physiologically realistic hemodynamic responses to accelerated atrial pacing. In practice, baseline MAP is used as a target: for each pacing rate, the activation timing is updated according to HR, and $C_{\mathrm{SV}}$ is optimized to maintain MAP. Optimization uses a combination of the genetic algorithm (GA) and *fminsearch* in MATLAB. The loss function is

$$\mathcal{L} = \left|\mathrm{MAP}_{\mathrm{paced}} - \mathrm{MAP}_{\mathrm{baseline}}\right|. \tag{9}$$

#### Estimation of Energetic Responses: LV $MVO_2$ and Cardiac Efficiency

While the hemodynamic response to atrial pacing is obtained directly from the digital twin model, the energetic response—including left-ventricular myocardial oxygen consumption (LV $MVO_2$) and cardiac efficiency (CE)—is not explicitly implemented in the model equations. Instead, we use the classical framework introduced by Suga and colleagues, in which myocardial oxygen consumption is strongly correlated with the pressure–volume area (PVA). PVA is defined as the sum of the external mechanical work (the area enclosed by the pressure–volume loop) and the potential energy, the area between the EDPVR and ESPVR [11].

Because the digital twin model directly computes instantaneous pressure and volume throughout the cardiac cycle, the pressure–volume loop is available without additional modeling. The remaining task is to estimate the end-diastolic pressure–volume relationship (EDPVR) and end-systolic pressure–volume relationship (ESPVR). Both are obtained using a single-beat estimation strategy.

The EDPVR is described by the exponential relationship [9]

$$P_{\mathrm{ED}}(V) = \beta\left[\exp\bigl(\alpha(V - V_0)\bigr) - 1\right], \tag{10}$$

where $\beta$, $\alpha$, and $V_0$ are parameters to be identified. To obtain multiple end-diastolic points for fitting, preload is perturbed by $-20\%$, $-10\%$, $+10\%$, and $+50\%$, generating five end-diastolic $(V_{\mathrm{ED}}, P_{\mathrm{ED}})$ pairs. Nonlinear least-squares fitting is then used to estimate $(\beta, \alpha, V_0)$.

Given $V_0$ from the EDPVR fit and the nearly linear nature of the ESPVR, the end-systolic pressure–volume relationship is obtained by connecting the end-systolic point during the simulated beat to $(V_0, 0)$:

$$P_{\mathrm{ES}}(V) \approx E_{\mathrm{es}}\,(V - V_0), \tag{11}$$

where $E_{\mathrm{es}}$ is the end-systolic elastance computed from the simulated end-systolic point.

Once EDPVR and ESPVR are identified, the total PVA is computed as

$$\mathrm{PVA} = \mathrm{EW} + \mathrm{PE}, \tag{12}$$

where EW is the external mechanical work (PV loop area) and PE is the potential energy enclosed between EDPVR and ESPVR.

Following the empirical relationship proposed by Takaoka et al [12], LV myocardial oxygen consumption per 100 g is calculated as

$$\mathrm{LVMVO_2} = \left(1.56 \times 10^{-5} \cdot \mathrm{PVA} + 0.00526 \cdot \frac{LV_\mathrm{m}}{100}\right) \cdot \mathrm{HR}, \tag{13}$$

where $LV_\mathrm{m}$ is the LV mass and HR is the heart rate.

Cardiac efficiency is computed as

$$\mathrm{CE} = \frac{\mathrm{CO}}{\mathrm{LVMVO_2}}, \tag{14}$$

where CO is the cardiac output simulated by the digital twin model.

#### Supplementary method 2: Details of The AI Model Development and Validation

##### Variational Autoencoder for Synthetic Digital-Twin Parameter Generation

A variational autoencoder (VAE) [13] was trained to generate physiologically plausible digital-twin (DT) model parameters. The model was implemented in Python using PyTorch and trained on the HFpEF-DT dataset, which contains 34 cardiovascular model parameters per subject.

Positive-valued variables were transformed using $\log_{10}$ to reduce right-skewness; nonpositive values were treated as `NaN`. Missing entries were imputed using a $k$-nearest-neighbor imputer ($k = 5$), although the final training dataset contained no missing values after preprocessing. Following imputation, all log-transformed variables were exponentiated ($10^x$) to return them to their original physical scale. Multiclass categorical variables were encoded using one-hot representation and then collapsed to index form for VAE input. All continuous variables were then scaled to the $[0, 1]$ interval using Min–Max normalization. After preprocessing, the resulting training tensor had dimensionality 34.

Features with $\leq 10$ unique values were treated as categorical; all others were treated as continuous. Let $d_\mathrm{cont}$ and $d_\mathrm{cat}^{(i)}$ denote the number of continuous variables and the number of categories in categorical variable $i$, respectively. The VAE outputs one Gaussian head for continuous variables and multiple categorical heads for discrete variables.

A fully connected encoder–decoder VAE was constructed with latent dimension 16. The encoder consisted of:

$$\mathbf{h}_1 = \mathrm{ReLU}(W_1\mathbf{x} + b_1), \qquad \mathbf{h}_2 = \mathrm{ReLU}(W_2\mathbf{h}_1 + b_2), \tag{15}$$

where $W_1 \in \mathbb{R}^{256\times34}$ and $W_2 \in \mathbb{R}^{16\times256}$. Two linear layers produced the latent mean and log-variance:

$$\boldsymbol{\mu} = W_\mu\mathbf{h}_2 + b_\mu, \qquad \log\boldsymbol{\sigma}^2 = W_\mathrm{log}\mathbf{h}_2 + b_\mathrm{log}. \tag{16}$$

Reparameterization was performed as

$$\mathbf{z} = \boldsymbol{\mu} + \boldsymbol{\sigma} \odot \boldsymbol{\epsilon}, \quad \boldsymbol{\epsilon} \sim \mathcal{N}(0, I). \tag{17}$$

The decoder applied:

$$\mathbf{h}'_1 = \mathrm{ReLU}(W'_1\mathbf{z} + b'_1), \qquad \mathbf{h}'_2 = \mathrm{ReLU}(W'_2\mathbf{h}'_1 + b'_2), \tag{18}$$

followed by two output branches:

- a linear layer for continuous variables ($d_\mathrm{cont}$ outputs),
- a set of categorical heads $\{W_c^{(i)} \in \mathbb{R}^{d_\mathrm{cat}^{(i)}\times128}\}$.

Categorical outputs were trained using raw logits; during sampling, categories were drawn using the Gumbel–Softmax distribution [14] with temperature $\tau = 0.5$.

For continuous variables, reconstruction error was computed as mean-squared error. For categorical variables, cross entropy was used. The total loss was:

$$\mathcal{L}_{\text{total}} = \mathcal{L}_{\text{recon}} + \mathcal{L}_{\text{cat}} + \lambda_{\text{KL}}\, D_{\text{KL}}\big(q_\phi(\mathbf{z} \mid \mathbf{x}) \parallel \mathcal{N}(0, I)\big)\,, \tag{19}$$

with $\lambda_{\text{KL}} = 5 \times 10^{-7}$. The KL term was computed as:

$$D_{\text{KL}} = -\frac{1}{2} \sum_{j=1}^{16} \left(1 + \log \sigma_j^2 - \mu_j^2 - \sigma_j^2\right). \tag{20}$$

The VAE was trained using the Adam optimizer (lr $= 3 \times 10^{-4}$, batch size 4) for up to 10,000 epochs with early stopping when the total loss fell below $8 \times 10^{-5}$. Training was performed on GPU when available.

To generate synthetic DT parameter sets, latent vectors were sampled from $\mathcal{N}(0, I)$ or from a covariance-preserving distribution using:

$$\mathbf{z} = \bar{\boldsymbol{\mu}} + L\,\boldsymbol{\epsilon}, \quad \boldsymbol{\epsilon} \sim \mathcal{N}(0, I), \tag{21}$$

where $L$ is the Cholesky decomposition of the empirical latent covariance. Decoded samples were post-processed by inverse Min–Max scaling and category selection via the Gumbel–Softmax sampler.

Each generated sample was passed to MATLAB through the Python–MATLAB interface and evaluated using the DT model. Only samples producing physiologically reasonable simulation were retained. A total of 30,000 samples were generated, of which 24,527 (75–85% acceptance) satisfied DT-model physiological consistency.To assess data diversity, all clinical variables in the real and generated cohorts were rounded to clinically relevant precision, and unique patient profiles were counted. After rounding, 100% of generated samples remained unique, indicating that the VAE decoder did not collapse to a limited set of clinical profiles.

#### Clinical-Feature Classifiers for Predicting Pacing Response

The virtual cohort generated from the VAE–digital-twin pipeline served as the training dataset for developing clinical-feature–based classifiers. The model was also implemented in Python using PyTorch. For each virtual subject, the digital twin provided simulated hemodynamic and energetic responses ($\Delta$LAP, $\Delta$SBP, $\Delta$CO, and $\Delta$CE) during accelerated atrial pacing. These responses were converted into binary labels as follows: Binary labels for the four pacing-response endpoints were assigned according to physiologically interpretable response directions. For $\Delta$LAP, a reduction in LAP was labeled as a favorable response (label = 1), whereas an increase was labeled as unfavorable (label = 0). For $\Delta$CO and $\Delta$CE, increases were labeled as favorable (label = 1) and decreases as unfavorable (label = 0). For $\Delta$SBP, a favorable hemodynamic response was defined as a systolic pressure reduction exceeding 8.5 mmHg (label = 1), with all smaller reductions or increases labeled as 0. The 8.5 mmHg cutoff was chosen solely to yield approximately balanced class distributions in the virtual cohort and does not imply clinical significance.

To enable clinical translation, only variables overlapping with routinely available measurements in the myPACE trial were used as classifier inputs. Seventeen features were included: sex, height, weight, systolic and diastolic blood pressure (SBP, DBP), ejection fraction (EF), septal and lateral left-ventricular wall thickness (`Hed_SW`, `Hed_LW`), heart rate (HR), left atrial maximum volume ($\text{LAV}_{\text{max}}$), early-to-late mitral inflow ratio (E/A), left-ventricular end-diastolic and end-systolic internal diameters (LVIDd, LVIDs), end-diastolic and end-systolic volumes (LVEDV, LVESV), pulmonary artery systolic pressure (PASP), and the presence of tricuspid regurgitation (TVr; yes/no). All features were normalized to the $[0, 1]$ interval using Min–Max scaling.

Four independent binary classifiers—one for each pacing-response endpoint (LAP, SBP, CO, CE)—were trained. Each classifier was implemented as a fully connected multilayer perceptron (MLP) with two hidden layers:

$$\text{MLP}: \quad 17 \;\rightarrow\; 32 \;\rightarrow\; 32 \;\rightarrow\; 1,$$

with ReLU activations in the hidden layers, dropout (0.5) between layers, and a sigmoid output unit. The model was trained using the Adam optimizer (learning rate $3 \times 10^{-4}$), batch size 48, and binary cross-entropy loss. Training was performed for up to 5,000 epochs until convergence.

The virtual cohort consisted of 24,527 subjects that passed physiological filtering by the digital-twin model and therefore served as the labeled training dataset. The dataset was randomly split into training (80%) and internal validation (20%) sets for each of the four binary classifiers. For each label, the optimal probability threshold was determined by maximizing the $F_\beta$ score (with $\beta = 1$ to prioritize precision), using the internal validation set.

To assess model generalization, the classifiers were evaluated on the HFpEF-DT cohort (146 real patients). Although these patients contributed to the distribution used to train the VAE, their digital-twin models were generated independently and were not part of the virtual cohort used for classifier training. The HFpEF-DT dataset provided physiologically simulated pacing responses, allowing measurement of AUROC, accuracy, and calibration without requiring clinical pacing data.

After threshold selection, the final classifiers were applied to the myPACE trial dataset. Missing values in myPACE were imputed using a $k$-nearest neighbors algorithm ($k = 5$), consistent with preprocessing in the training pipeline. Since myPACE lacks real pacing-response measurements ($\Delta$LAP, $\Delta$SBP, $\Delta$CO, and $\Delta$CE), classifier outputs were used as real-world predictions of favorable vs. unfavorable hemodynamic and energetic responses to atrial pacing.
